# Deep brain stimulation and psychosis: A case series and two candidate causal brain circuits

**DOI:** 10.1101/2025.11.18.25340443

**Authors:** Garance M. Meyer, Andrew R. Pines, Alexandra Roldan, Ignacio Aracil-Bolaños, Ella Gray Settle, Ilkem Aysu Sahin, Frederic L. W. V. J. Schaper, Clemens Neudorfer, Soila Järvenpää, Hikaru Kamo, Sylvie H. M. J. Piacentini, Luigi M. Romito, Min Jae Kim, Lukas L. Goede, Konstantin Butenko, Bahne H. Bahners, Philip E. Mosley, Alexandre Rainha Campos, Karmele Olaciregui Dague, James Boyd, Alik S. Widge, Darin D. Dougherty, Yasin Temel, Rob P.W. Rouhl, Albert Colon, Andrea Kühn, Mohammad Maarouf, Antonella Macerollo, Jibril Osman Farah, Genko Oyama, Nobutaka Hattori, Kai Lehtimäki, Jukka Peltola, Harith Akram, Thomas Foltynie, Chencheng Zhang, David Silbersweig, Michael D. Fox, John D Rolston, Juan Angel Aibar-Durán, Iluminada Corripio, Shan H. Siddiqi, Andreas Horn

## Abstract

Schizophrenia and psychosis are debilitating conditions with suboptimal treatment options. Deep brain stimulation (DBS) offers promise, but effective treatment targets remain undefined. Examining cases in which DBS either induced or alleviated psychotic symptoms may help identify circuits causally involved in psychosis and suggest candidate targets for intervention.

We systematically reviewed the literature to identify all published cases in which DBS modulated (i.e., caused or improved) psychotic symptoms, regardless of target and indication. Authors of original publications were contacted to gather individual case data, allowing DBS electrode reconstruction and stimulation volume modeling. This data was aggregated into standard space and used to characterize anatomical structures most consistently associated with change in symptoms.

After screening 332 studies, 36 cases were retained. This included 16 patients who received DBS for treatment-resistant schizophrenia or psychosis (nucleus accumbens, N=7; subgenual cingulate, N=4; substantia nigra pars reticulata, N=3; habenula, N=2) and 18 patients who received DBS for treatment of other conditions and experienced psychotic symptoms as a side effect (anterior nucleus of the thalamus, N=7; centromedian nucleus, N=1; subthalamic nucleus, N=6; nucleus accumbens, N=2; globus pallidus pars interna, N=1; amygdala, N=1). Finally, DBS of the nucleus basalis of Meynert improved visual hallucinations in two additional cases. Although stimulation sites were anatomically heterogeneous, qualitative integration of the empirical anatomical findings with current neurobiological models of schizophrenia revealed two circuits potentially implicated in psychotic symptoms: one centered on the mediodorsal nucleus of the thalamus and its main subcortical afferents, and one involving the nucleus accumbens – ventral tegmental area loop.

We propose a preliminary theoretical framework linking these circuits to the emergence and improvement of psychotic symptoms, thereby generating testable hypotheses for future mechanistic and clinical studies. We suggest that disruption of these circuits may respectively relate to impaired filtering of cognitive and limbic representations, and aberrant salience processing.

## 1 Introduction

Deep brain stimulation (DBS) is an established treatment for movement disorders and epilepsy.^1^ In recent years, it has been increasingly explored as a therapeutic option for treatment-resistant psychiatric disorders^2^, including schizophrenia, for which the first contemporary clinical trials and case reports have been published in the last five years.^3–7^ Indeed, up to 20-30% of patients with schizophrenia experience persistent psychotic symptoms—including delusions and hallucinations—despite adequate pharmacological treatment^8–10^, and many experience intolerable side effects from these drugs^11,12^. For these reasons, there has been growing interest in neuromodulation as a treatment option^13^. While numerous brain network abnormalities have been reported in schizophrenia^14^, the causal mechanisms of the disease are not fully identified. Hence, the first DBS trials explored a variety of surgical targets^13,15–17^; included in Results below), but the best candidates and overall suitability of DBS remain uncertain.

Over the years and across indications, retrospective studies of anatomical correlates of treatment response^18–22^ and serendipitous clinical observations^23–26^ have played a key role in defining and refining DBS targets. Here, we build on both approaches to investigate potential DBS targets for treatment-resistant schizophrenia. Importantly, experience from other indications suggests that defining an anatomical region or nucleus as the DBS target may be insufficient; instead, defining precise targeting coordinates or sweet-spots *within* such targets is essential.^20,22^

We assembled the largest DBS case series to date in which psychotic symptoms were affected by stimulation, either intentionally or incidentally (N=36). In other words, we included cases of DBS for schizophrenia and cases in which psychotic symptoms serendipitously emerged or improved in patients receiving DBS for other indications. The purpose of the study was two-fold. First, we provide an objective, quantitative characterization of the anatomical structures engaged by stimulation in these cases. To this aim, original imaging data was retrieved, and electrodes were localized for 29 cases, through collaboration across 17 institutions worldwide. Stimulation volumes were modeled and their overlap with surrounding structures was calculated. Second, we interpret these anatomical findings in the context of existing models of schizophrenia^13–17^, hypothesizing that stimulation sites causing or improving psychotic symptoms would map onto a limited number of brain circuits implicated in these models. We propose a preliminary theoretical framework linking specific circuits to the emergence and improvement of psychotic symptoms, thereby generating testable hypotheses that may guide future mechanistic and clinical studies.

## 2 Methods

### 2.1 Literature search

We performed a systematic review of the literature to identify reported cases in which deep brain stimulation modulated psychotic symptoms such as auditory or visual hallucinations and/or delusions, irrespective of surgical target and indication. We included patients receiving DBS for treatment-resistant psychosis, as well as patients who received DBS for different indications but presented with new onset or improved psychotic symptoms following stimulation. Importantly, for incidental cases, we only included cases with evidence of causal relationship between stimulation and change in symptoms. For example, we included cases in which symptoms started shortly after a change in stimulation settings and improved upon further change in settings or discontinuation of stimulation, but not cases in which symptoms were already present before surgery and worsened or re-occurred under DBS. Authors of suitable publications were contacted by the team at Mass General Brigham to gather individual case data. Detail about case selection is available as Supplementary Methods.

Written informed consent was initially obtained from all patients by corresponding centers. The present study (secondary data analysis) was approved by the local Institutional Review Board in accordance with the Declaration of Helsinki.

### 2.2 Anatomical analysis

When possible, the electrodes were reconstructed using the standard pipeline of the Lead-DBS toolbox^27^ based on the preoperative and postoperative imaging. In some cases, approximative electrode reconstructions were based on imaging shown in the respective publications (Supplementary Methods). Stimulation volumes were modeled based on available stimulation parameters using the OSS-DBS software.^28^

To identify the stimulated white matter pathways and structures, the volumetric overlap between the stimulation volumes and surrounding structures was computed on the basis of anatomical atlases and normative connectomes available in Lead-DBS.^29–33^ Additionally, to account for subcortical pathways that are typically absent or misrepresented in connectomes derived from diffusion-weighted imaging^34^, we included anatomically defined tracts from the FOCUS atlas.^35,36^

Anatomical findings were interpreted in the context of current neurobiological theories of schizophrenia^13,15–17^ to identify candidate circuits consistently modulated across cases. Relevant theoretical models and corresponding circuits were identified through literature review and are summarized in Table S1. Given the small and heterogeneous sample, this step of the analysis was qualitative and exploratory, aiming at integrating the empirical anatomical findings with existing models, rather than performing data-driven statistical testing.

## 3 Results

Thirty-four relevant cases were identified from the literature, and two additional cases were shared by co-authors, leading to the inclusion of 36 patients in this case series. Electrode reconstructions could be performed for 29 of these cases (Fig. 1; see Fig. S1 for a flow chart). Clinical information is available in Table S2 and S3. Detailed anatomical results (i.e., volumetric overlap results) are presented in Figures S2-S11 (featuring 2-D views) and Tables S4-S14.

**Figure 1:**
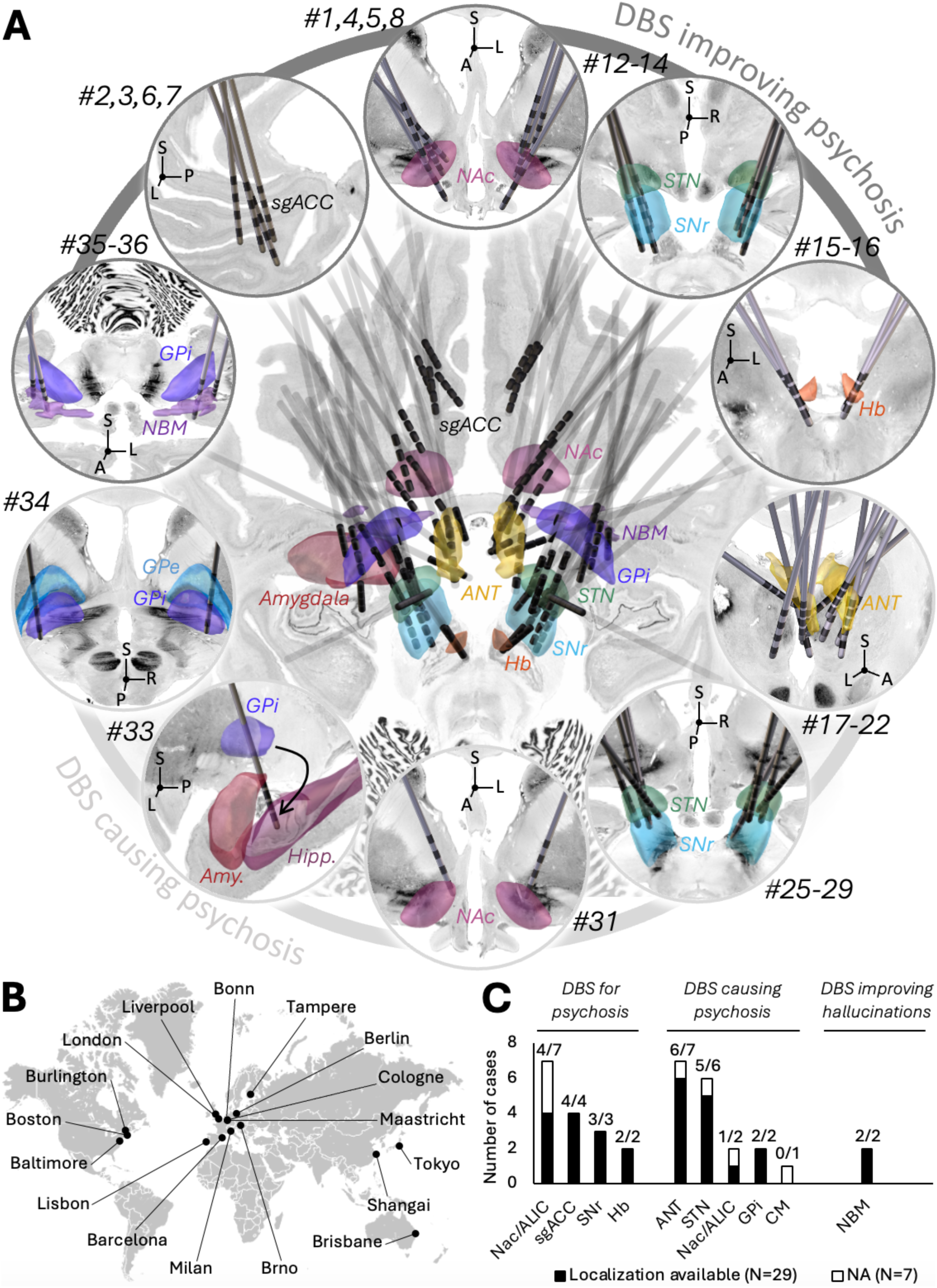
Overview of cases of DBS causing or improving psychosis. (A) Electrode positions for the cases with available electrode reconstructions (N=29 out of 36 total cases retrieved from the literature). (B) These electrode reconstructions were gathered through collaboration between 17 institutions worldwide (map from www.openstreetmap.org). (C) Break down of all 36 cases per anatomical target. DBS for treatment-resistant schizophrenia or psychosis has been attempted at the subgenual cingulate (sgACC, cases #2,3,6,7), nucleus accumbens (NAc, cases #1,4,5,8), substantia nigra pars reticulata (SNr, case #12-14), and habenula (Hb, cases #15-16). DBS triggered psychotic symptoms when applied at various targets and for various indications, including the anterior nucleus of the thalamus in patients with epilepsy (ANT, cases #17-22), the subthalamic nucleus in patients with PD (STN, cases #25-29), the nucleus accumbens in a patient with OCD (case #31), and the globus pallidus pars interna in a patient with PD (GPi, case #34). In an additional case, a patient with dystonia developed psychosis after the left GPi electrode migrated to the amygdala region (case #33). Finally, two patients with PD dementia reported improved visual hallucinations under DBS of the nucleus basalis of Meynert (NBM, cases #35-36). Electrodes are shown against 3-D structures from the DISTAL^29^, CIT-168^32^, Morel atlas^37^, and atlas of the human hypothalamus^31^, overlaid on the Big Brain template^38,39^, postero-superior view). GPe: globus pallidus pars externa, Hipp.: hippocampus.

### 3.1 DBS for treatment of schizophrenia and psychosis

Sixteen patients received DBS for treatment-resistant schizophrenia or secondary psychosis (Cases #1-16, 13 with available electrode reconstructions, Table S2; Fig. 2).

**Figure 2:**
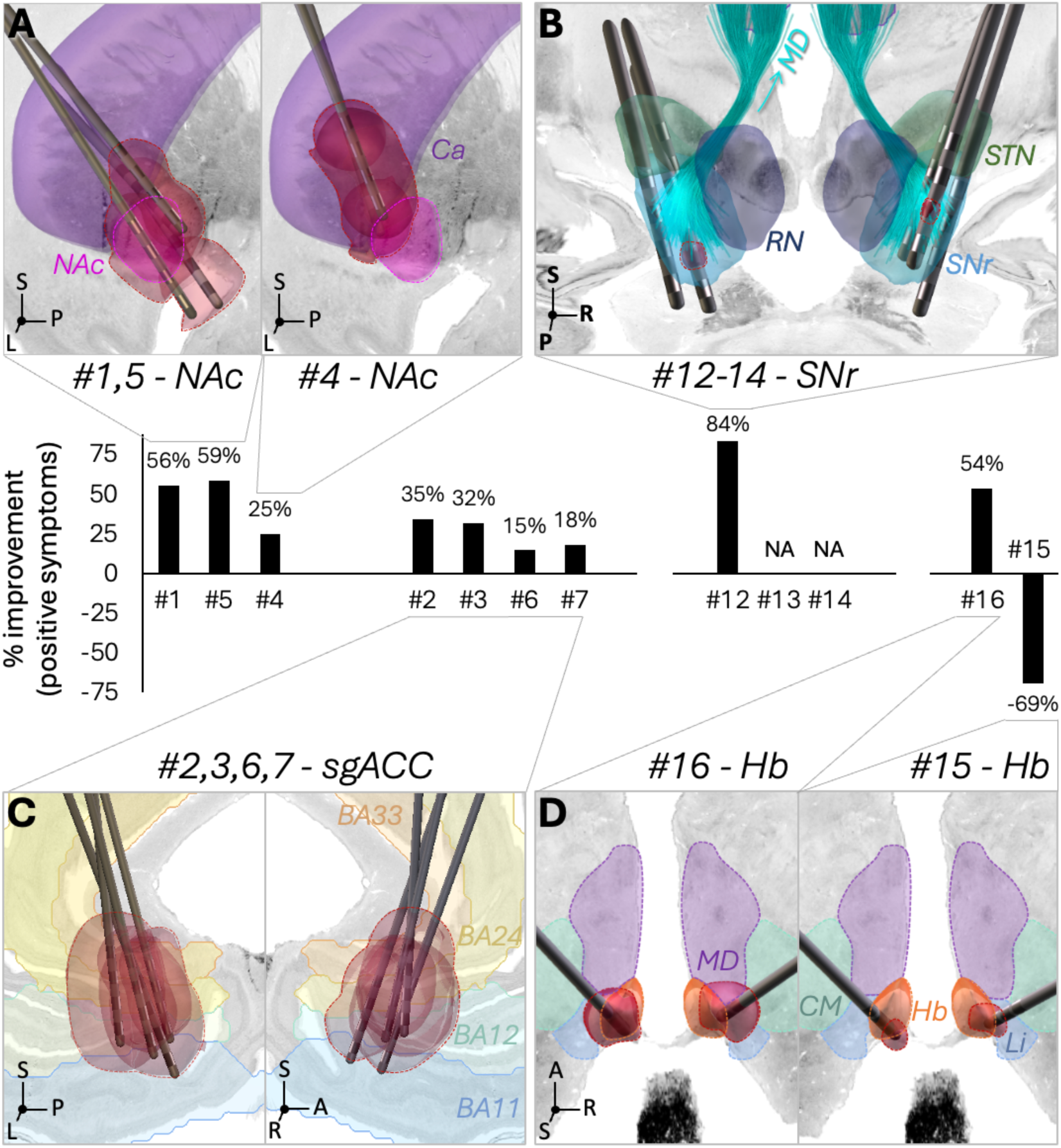
Deep brain stimulation for schizophrenia. (Cases #1-16). (A) Six patients received NAc-DBS. Only three patients with available stimulation volumes are represented (stimulation volumes in red); the two patients who improved the most had stimulation volumes mostly covering the NAc and surrounding limbic pathways (Cases #1,5), while the patient who improved the least was mostly stimulated in the caudate (Case #4). (B) SNr-DBS was performed in three patients (Cases #12-14). Chronic stimulation parameters and clinical outcomes were only available for one patient, who improved by 84% (stimulation volumes in red). All electrodes had contacts in the SNr. The nigrothalamic projections to the mediodorsal nucleus of the thalamus are represented, as indicated by the cyan arrow. (C) sgACC-DBS was performed in four patients (Cases #2,3,6,7; left and right hemispheres shown separately). There was no obvious relationship between stimulation location and outcomes. (D) Two patients received DBS of the habenula. One patient improved (Case #16) while the other worsened (Case #15); the patient who improved had larger stimulation volumes (red), resulting in more extensive coverage of the habenula and surrounding thalamic nuclei. (D) BA: Brodmann area, Ca: caudate, CM: centromedian nucleus of the thalamus, Hb: habenula, Li: limitans nucleus, MD: mediodorsal nucleus of the thalamus, NAc: nucleus accumbens, RN: red nucleus, SNr: substantia nigra pars reticulata, STN: subthalamic nucleus.

Six patients—including a patient with schizoaffective disorder—received NAc-DBS, with a mean improvement of 28.2% on the Positive and Negative Syndrome Scale (PANSS; 46.5% on the positive symptoms sub-score for the three patients for whom this information was available). One patient entered a double-blind crossover phase (ON vs. OFF stimulation) and demonstrated worsened symptoms while OFF stimulation. Two patients experienced incidental deactivation of the device, resulting in rapid worsening of symptoms for one patient and no discernible effect for the other.

Four patients received DBS of the subgenual cingulate (sgACC; Cases #2,3,6,7) and experienced a mean improvement of 19.6% on the PANSS (24.8% on the positive symptoms sub-score). Two of these patients entered a double-blind crossover phase and both experienced worsened symptoms while OFF stimulation.

Electrode reconstructions were available for four NAc-DBS (including one with unknown stimulation parameters, for whom stimulation volumes could not be calculated) and four sgACC-DBS cases (Cases #1-8). While robust relationships between treatment outcomes and stimulated anatomical structures could not be statistically explored in such a small sample, the two top responders received NAc- (rather than sgACC-) DBS, with stimulation volumes mostly covering the NAc and surrounding limbic pathways (including dopaminergic projections from the VTA). In contrast, the third patient with NAc-DBS and available stimulation parameters, who demonstrated a lesser response to stimulation and no change in symptoms during deactivation of the device, had predominant caudate stimulation and lesser engagement of the above-mentioned white matter pathways (despite larger stimulation volumes). No obvious relationship between anatomical substrates and outcomes could be derived for the patients with sgACC-DBS (Fig. S2, Table S4 and S5).

Two patients received DBS of the habenula (Cases #15,16). One of these patients experienced a 31.7% improvement on the PANSS (53.8% on the positive symptoms sub-score), while the other one worsened by 9.5% (69% for positive symptoms) and was withdrawn from the study after 10 months. The patient who improved had larger stimulation volumes, resulting in more extensive coverage of the habenula, as well as, notably, of surrounding thalamic nuclei such as the central lateral, central medial and mediodorsal (MD) nuclei (Fig. S4, Table S6).

Three patients received DBS of the SNr (Cases #12-14). To date, clinical outcomes have only been reported for one of them. This patient experienced acute, reproducible resolution of hallucinations, as well as an 84% improvement in Brief Psychotic Rating Scale (BPRS) scores for unusual thought content, hallucinations and delusional suspiciousness at 6 months. Stimulation volumes covered the posterior aspect of the SNr (Fig. S3). In the two other patients, all electrodes had at least two contacts in the SNr (Fig. 2D).

Finally, a patient with psychosis secondary to traumatic brain injury received DBS of the NAc/ALIC (Case #11, electrode reconstruction not available), which resulted in a 64.5% improvement on the PANSS.

### 3.2 Incidental cases

We identified 20 cases in which DBS incidentally triggered (N=18) or improved (N=2) psychotic symptoms (Table S3).

#### 3.2.1 ANT- and CM-DBS

Seven of the incidental cases involved patients with epilepsy receiving DBS of the anterior nucleus of the thalamus (ANT; Cases #17-23; 6/7 with available electrode reconstructions). The patients all presented with delusions, and symptoms could be relieved by DBS reprogramming in five cases (two reduced voltage, three switched to more dorsal contacts). In all 6 cases with available electrode reconstructions, stimulation volumes that induced psychotic symptoms overlapped with the ANT and MD, albeit to varying degrees. Overlap with surrounding thalamic nuclei was also seen, such as the central lateral (6/6 patients) and ventrolateral posterior (5/6 patients) nuclei. Interestingly, reprogramming led to numerical decrease in stimulation overlap with the MD in all four patients with available electrode reconstruction who underwent successful reprogramming, with concomitant *increase* in overlap with ANT in 3/4 (Fig. 3A-D, Fig. S5, Table S7 and S8).

**Figure 3:**
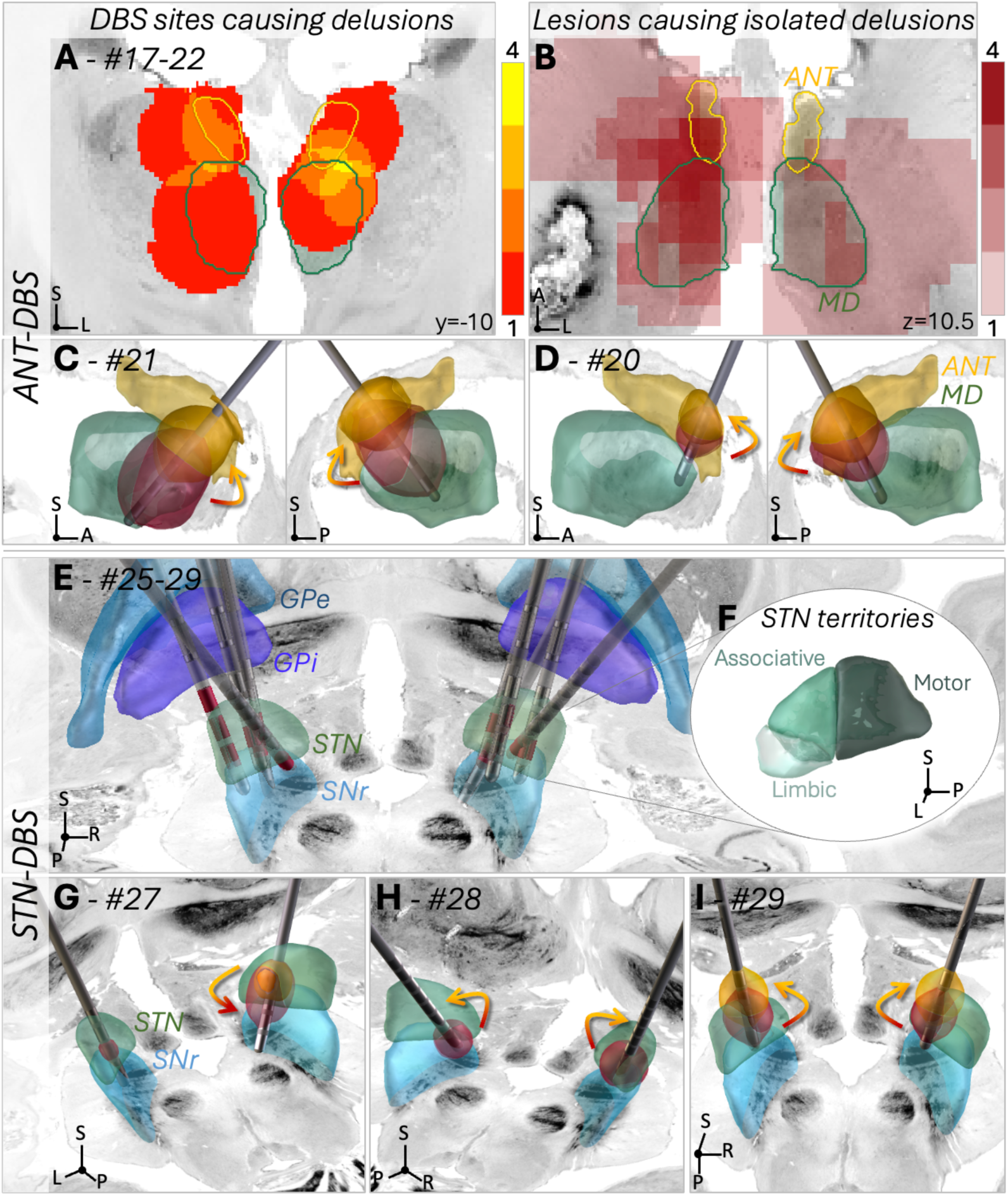
Cases of psychosis in patients receiving ANT- or STN-DBS. (Cases #17-22 and #25-29). (A) Cases #17-22 – Seven patients with epilepsy receiving ANT-DBS developed psychosis. The patients all presented with delusions, and symptoms could be relieved by reprogramming of the stimulation in five cases. Stimulation volumes overlapped with both the ANT and mediodorsal nucleus of the thalamus (MD) in each of the six cases with available electrode reconstructions. A N-map of stimulation volumes is shown. (B) Consistent with these observations, lesions of the MD have been reported to cause isolated delusions (seven out of the eight lesion cases with isolated persecutory delusions reported in^40^ intersected with the MD; a N-map is shown;^41–46^). (C,D) Example patients in whom reprogramming of the stimulation to more dorsal contacts (indicated by the arrows), resulting in decreased involvement of the MD, resolved psychotic symptoms. 2-D views of all stimulation volumes are available as Fig. S5. (E) Cases #25-29 – These five patients with PD experienced psychotic symptoms during STN-DBS. Active contacts are highlighted in red. 2-D views of all stimulation volumes are available as Fig. S7. (F) Functional territories of the subthalamic nucleus^47,48^. (G) Case #27 developed persecutory delusions and manic symptoms after activation of an additional ventral contact, which resulted in stronger involvement of the SNr (yellow: stimulation volumes before reprogramming, red: stimulation volumes associated with symptom onset). (H) Case #28 developed psychosis and manic symptoms upon initiation of stimulation, which subsided when stimulation was switched to more dorsal contacts (not shown, precise stimulation parameters not available), likely resulting in reduced involvement of the SNr and limbic STN. (I) Case #29 developed psychosis after an increase in stimulation amplitude (red stimulation volumes). Symptoms resolved with reprogramming to more dorsal contacts (yellow stimulation volumes), which resulted in reduced involvement of the limbic STN and SNr. Arrows indicate DBS reprogramming. ANT: anterior nucleus of the thalamus, GPe: globus pallidus pars externa, GPi: globus pallidus pars interna, MD: mediodorsal nucleus of the thalamus, SNr: substantia nigra pars reticulata, STN: subthalamic nucleus.

One patient with epilepsy developed psychosis under DBS of the centromedian (CM) nucleus of the thalamus, characterized by auditory hallucinations, which improved upon reprogramming (Case #24, electrode reconstructions not available). Given that we found no other published cases of psychosis after CM-DBS, these symptoms may be hypothesized to result from off-target stimulation effects on adjacent thalamic structures, potentially the MD or pulvinar (see Fig. S6 for a representative CM electrode).

#### 3.2.2 STN-DBS

Six cases involved patients with Parkinson’s disease (PD) receiving DBS of the subthalamic nucleus (STN; Case #25-30; 5/6 with available electrode reconstructions). Psychotic symptoms developed in the context of mania in three of these six cases (Cases #27, #28, and #30). Of note, it is well known that STN-DBS for PD sometimes causes hypomania in patients with no significant psychiatric history, which is typically improved by change in stimulation parameters ^49,50^. Here, we included only cases in which clear psychotic symptoms –such as delusions or hallucinations– were present, beyond the grandiosity that may occur in hypomania or mania.

Stimulation volumes overlapped with the limbic territory of the STN in all five patients with available electrode reconstructions, and with SNr in 4/5. In two cases, symptoms improved when switching to more dorsal contacts, and in one case, they only started after adding a more ventral contact. Anatomically, stimulation at the more ventral contacts resulted in stronger overlap of stimulation volumes with SNr and limbic STN, as well as in stronger involvement of nigrothalamic efferents to MD and limbic portion of the hyperdirect pathway (Fig. 3E-H, Table S9 and S10; Fig. S7).

#### 3.2.3 NAc-DBS

Delusions occurred after DBS of the NAc region in two cases (one major depressive disorder, one obsessive compulsive disorder, Cases #31 and #32), without any signs of mania, and could be relieved by reprogramming or discontinuation of the stimulation. For the patient with available electrode reconstruction (#31), stronger involvement of the anterior commissure, stria terminalis, and limbic portion of the hyperdirect pathway was observed for stimulation settings linked to the emergence of symptoms (Fig. S8, Table S11).

#### 3.2.4 GPi-DBS

Two patients developed psychosis after DBS of the globus pallidus pars interna (GPi; one PD, one dystonia). The first patient (Case #34) experienced acute severe paranoia, depression, and suicidality, which could be attributed to stimulation of contacts dorsal to the GPi on the right electrode, and could be resolved by stimulating the more ventral right contacts within the GPi (Fig. S10 and Table S13). Interestingly, a lesion could be seen on the MRI (hyperintense on the T2w image) within the white matter medial to the globus pallidus (internal capsule and pallidothalamic projections, including projections from GPi to MD). Critically, the lesion was closest to the contact that first elicited symptoms. The presence of this lesion may have distorted the current flow, leading to shunted current and unexpected stimulation effects at a distance from the stimulating contact (Fig. 4A-C)

**Figure 4:**
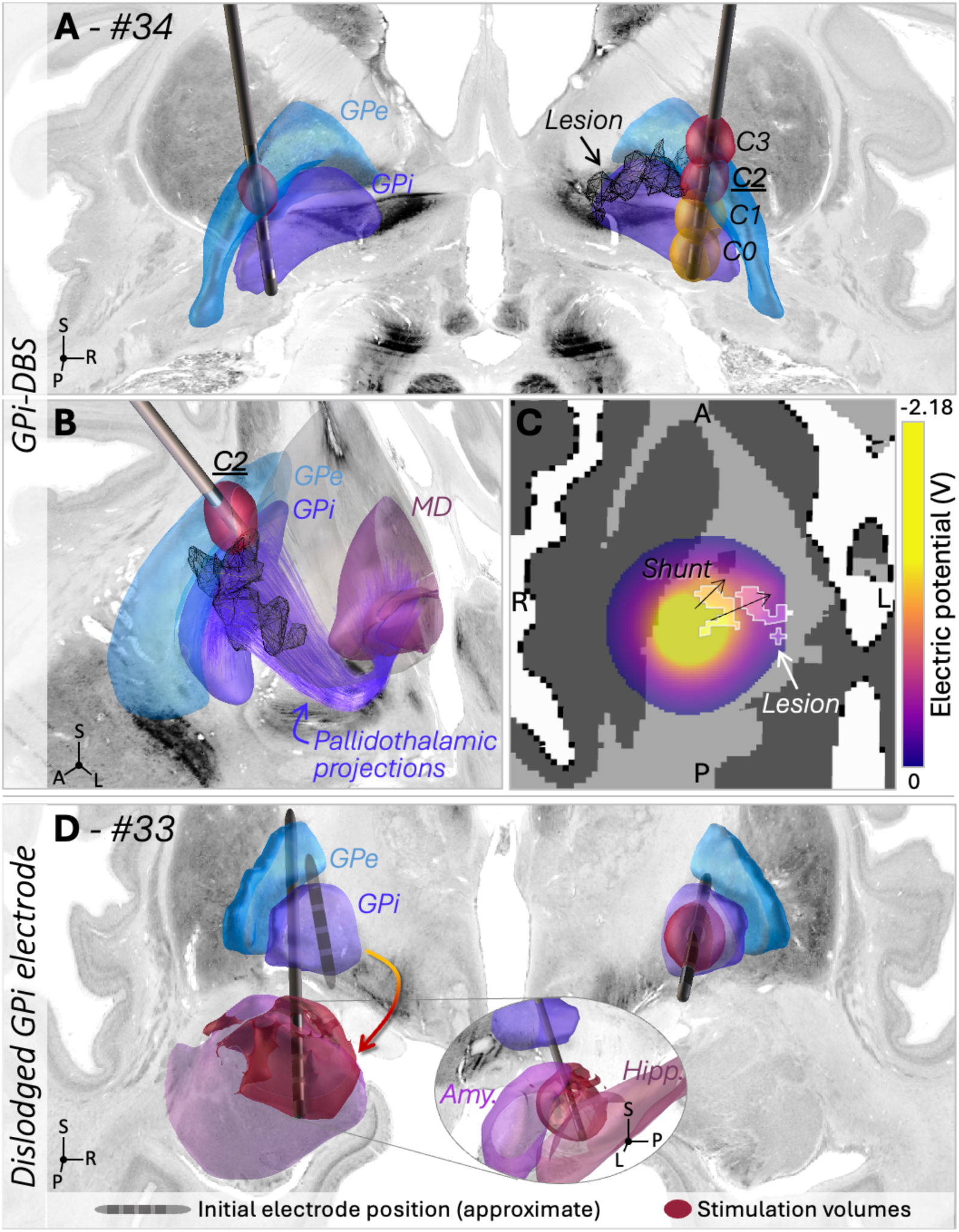
Cases of psychosis in patients receiving GPi-DBS. (Cases #33 and 34; posterior view). (A) Case #34 – This patient with PD developed severe paranoia, depression and suicidality within a few hours of starting stimulation at the third contacts on each side (C2). Symptoms ceased abruptly when discontinuing stimulation. Further attempts to use right contacts C2 and C3 (red stimulation volumes), but not C0 and C1 (yellow stimulation volumes), led to the recurrence of symptoms. Additional 2-D views of stimulation volumes are available as Fig. S10. (B) The T2w MRI showed a hyperintense lesion in the vicinity of right contact C2, which overlapped with surrounding white matter, including pallidothalamic projections (purple) and internal capsule (passing between the thalamus and globus pallidus; not shown). (C) While the impact of the lesion cannot be modeled with certainty, E-field modeling using the OSS-DBS toolbox^28^ suggests that its presence distorted the current flow, potentially leading to unexpected stimulation effects at a distance from the stimulating contact, in the abovementioned white matter. (D) Case #33 – This patient with dystonia developed marked psychotic symptoms 5-9 months after implantation, which was attributed to the left electrode having migrated from its original position (approximate location shown as a schematic electrode) to the amygdala region, as represented by the arrow. As a result, the stimulation volume covered parts of the amygdala and hippocampus. Symptoms were relieved by discontinuation of the stimulation and electrode repositioning. Additional 2-D views of stimulation volumes are available as Fig. S9. GPe: Globus pallidus pars externa, GPi: Globus pallidus pars interna, MD: Mediodorsal nucleus of the thalamus.

In the second case (Case #33), a patient with no psychiatric history presented with delusions, mood lability, and depression. The clinical team discovered the left electrode had migrated, with the corresponding stimulation volume covering parts of the amygdala and hippocampus (Fig. 4D, Fig. S9, and Table S12). Symptoms were relieved by discontinuation of the stimulation and left electrode repositioning.

#### 3.2.5 NBM-DBS

In the last two cases, DBS of the nucleus basalis of Meynert (NBM) applied to treat PD dementia caused near-complete cessation of pre-existing visual hallucinations. Hallucinations recurred when stimulation was temporarily turned off (Cases #35 and #36). These cases were characterized by more antero-ventral electrode placement as well as stronger involvement of surrounding white matter pathways (anterior commissure, ventral amygdalofugal pathway) as compared to two patients from the same trial whose pre-existing hallucinations did not improve. The degree of overlap with NBM was similar across cases^a^ (Fig. S11, Table S14).

### 3.3 Mapping anatomical findings to two candidate circuits involved in psychosis

In a qualitative synthesis, most of the cases presented here involved stimulation sites modulating two candidate circuits previously implicated in psychosis (see Table S1 for a summary of previous evidence). Six cases could be mapped to the first circuit, consisting of the dopaminergic VTA-NAc loop and its hippocampal input, and 20 cases could be mapped to the second one, which was centered on the MD and its main subcortical afferents (i.e., amygdala and SNr/GPi; see Discussion for contextualization with known neuroanatomy). A summary of findings is presented in Fig. 5.

**Figure 5:**
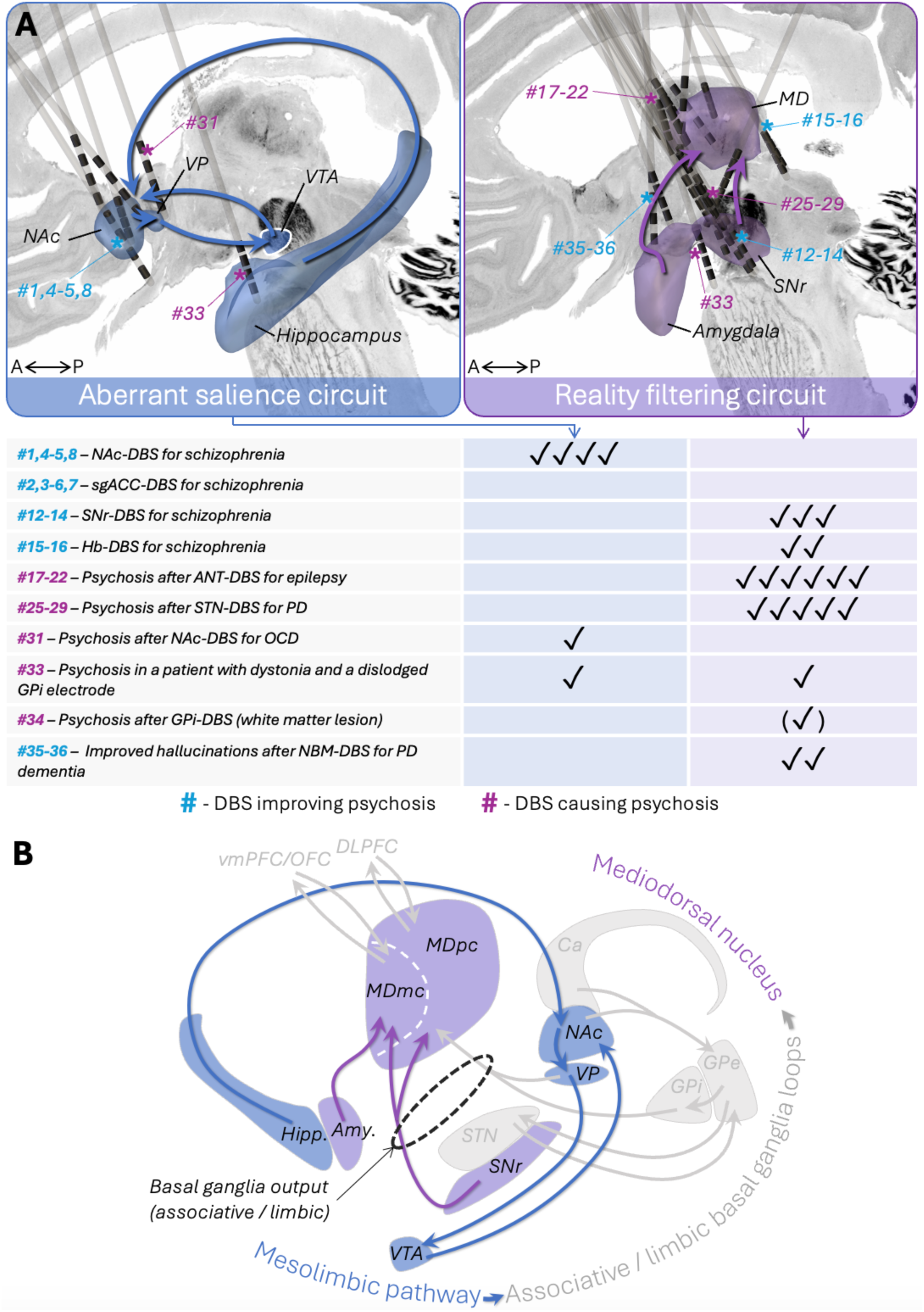
Summary of findings and a proposed preliminary theoretical framework. Across all cases with available electrode reconstructions (N=29), stimulation sites converged on two candidate circuits previously implicated in schizophrenia. First, six cases could be tied to the involvement of the dopaminergic VTA-NAc loop and its hippocampal input. Specifically, NAc-DBS was more effective for the treatment of psychotic symptoms than sgACC-DBS in the Barcelona cohort (Cases #1-7), and, across the patients with NAc-DBS, better outcomes were observed for patients with stronger involvement of NAc and dopaminergic projections from the VTA (Cases #1,5) rather than caudate nucleus (Case #4). Psychotic symptoms also appeared under NAc-DBS (Case #31), and under stimulation of the hippocampus and amygdala through a dislodged electrode (Case #33). Second, twenty cases could be mapped to the MD or its main subcortical afferents (i.e., projections from the amygdala and GPi/SNr). Consistent with the effects of MD lesions, cases of psychosis after ANT-DBS involved MD stimulation in all cases and were relieved by reprogramming decreasing MD involvement (Cases #17-22). Third, psychosis after STN-DBS (Cases #25-29) was associated with involvement of the SNr (also consistent with a previous lesion case^54^). On the other hand, SNr-DBS was effective for the treatment of schizophrenia (Cases #12-14). Three other cases implicated projections from the amygdala to MD or the amygdala itself, including the two patients who experienced improved hallucinations with NBM-DBS (Cases #35-36), and the patient who developed psychosis after electrode displacement to the amygdala region (Case #33). Finally, improvement of psychotic symptoms with habenular DBS was associated with stimulation volumes encompassing the MD (Cases #15-16), and the patient who developed psychosis under GPi-DBS had a lesion neighboring the stimulated contact, which may have resulted in unintended shunting of current toward projections from GPi to MD (Case #34). Pink and blue stars represent cases in which DBS caused and improved psychosis, respectively. We propose that disruption of these two circuits may respectively relate to aberrant salience processing, and impaired filtering of cognitive and limbic representations (see Discussion for detail). (B) Of note, these two candidate circuits may be seen as subcomponents of a larger overarching loop. Indeed, the mesolimbic pathway (VTA to NAc) may be seen as an entry point of the limbic basal-ganglia thalamocortical loop, while the MD constitutes a key convergence point for both the limbic (magnocellular part; MDmc) and associative loops (parvocellular part; MDpc), as the main thalamic nucleus receiving limbic and associative output from the basal ganglia through nigrothalamic (SNr → MD) and pallidothalamic projections (GPi → MD and VP → MD)^55^.

## 4 Discussion

Our case series describes the largest collection to date of cases in which DBS modulated psychotic symptoms (N=36). This includes contemporary cases of DBS for treatment-resistant schizophrenia or psychosis (see Supplementary material 2 for an account of early attempts in the 1950s^51–53^), and cases in which psychotic symptoms emerged or improved in patients receiving DBS for other indications. Such cases are exceedingly rare and could only be gathered through extensive literature screening and multicenter collaboration. First, we performed detailed characterization of the anatomical structures engaged by stimulation in 29 cases with available electrode reconstructions. Of note, emergence of psychotic symptoms could be linked to off-target DBS effects in a majority of cases. Second, by qualitatively integrating our anatomical findings with current neurobiological models of schizophrenia, we identified two candidate circuits that may be causally involved in psychotic symptoms and might constitute promising targets for DBS (Fig. 5). The potential functional significance of these circuits is discussed below.

The first candidate circuit consists of mesolimbic dopaminergic projections from the VTA to NAc, and feedback inhibitory projections from the NAc to VP and VP to VTA, which in turn participates in the regulation of dopamine release in the NAc. Six of our cases could be tied to modulation of this circuit (see Fig. 5), which has been consistently implicated in psychosis. Indeed, a hyperdopaminergic state in the striatum is a consistent finding in studies of schizophrenia (dopamine hypothesis of schizophrenia^56,57^; Table S1). Hippocampal dysfunction is another consistent finding^58,59^; Table S1) and has been proposed to drive the dysregulation of mesolimbic dopamine transmission by interfering with the feedback regulation of dopamine release from the VTA^60^. Mechanistically, the hyperdopaminergic state resulting from disruption of this circuit is believed to result in aberrant salience signaling^61^. Aberrant assignment of salience to internal or external stimuli –such as voices, sounds, or thoughts– may result in hallucinations and delusions as “top-down” cognitive narratives imposed on these perceptions in an effort to provide explanatory frameworks for their salience^61,62^. Of note, the effects of NAc-DBS on dopaminergic transmission remains unclear and cannot be inferred from the present study^16,17,63^.

The second candidate circuit is centered on the medio-dorsal nucleus of the thalamus, including projections from the SNr and amygdala, which constitute two of its main subcortical afferents^55,64–66^. Twenty cases could be mapped to this circuit (Fig. 5). Numerous lines of evidence implicate the MD in psychosis (see Table S1), including reduced MD volume in schizophrenia (neuroimaging and post-mortem studies^67^), consistent alterations of thalamocortical connectivity –particularly MD-PFC connectivity^67,68^, and the occurrence of delusions with MD lesions^40^ (Fig. 3B). Despite this converging evidence, and in contrast to the VTA-NAc loop, no comparably strong theoretical model has been proposed to link MD dysfunction to symptoms of psychosis. Several observations allow us to speculate on exactly this aspect. First, the MD—together with the ventral anterior nucleus, to a lesser extent—provides the sole thalamic relay for basal ganglia output in associative and limbic cortico–basal ganglia–thalamocortical loops, through nigrothalamic and pallidothalamic projections^55,69^. Specifically, its lateral, parvocellular part, connects with the DLPFC (associative loop), while its medial, magnocellular part receives dopaminergic innervation from the SNr and VTA, connects with the OFC, and integrates amygdala input via the amygdalofugal pathway (limbic loop).^55,69^ Computational accounts of thalamic function suggests that the role of the MD may be summarized as monitoring, maintaining and updating relevant cognitive (associative) and affective (limbic) representations^70–74^, consistent with its involvement in a variety of cognitive processes such as memory and executive functions.^75–79^ In line with this overarching computational function of the MD, the amygdala-MD-OFC loop has been implicated in a cognitive process that bears direct relevance for psychotic symptoms, namely, reality filtering, or the process by which representations that are not relevant for ongoing reality are suppressed, and relevant representations selected.^80–84^ Amygdala input may be crucial for this process, potentially flagging the emotional (i.e., threat) significance of these representations.^85,86^ We propose that MD dysfunction or disruption interferes with reality filtering, leading to inappropriate maintenance of irrelevant representations and hereby contributing to hallucinations and delusions. Disruption of amygdala input may specifically lead to the attribution of erroneous threat salience to irrelevant representations, potentially contributing to persecutory and paranoid delusions^87^, which were consistently observed in our ANT-DBS cases (Cases #17-22).

It remains unclear whether psychotic symptoms can arise from disruption of any of these two circuits or require combined disruption, as proposed in “two-hit” models.^88,89^ Essentially, the “two-hit” hypotheses suggest that psychotic symptoms result from the simultaneous introduction of noisy information in the system (for example, because of altered visual inputs^88^; or because of aberrant salience attribution^57^) and disruption of the ability to filter out this noisy information—a function that may be supported by the MD, as detailed above. In some of the presented cases, disruption of both circuits may have occurred. For example, in Case #33, both the amygdala and hippocampus were stimulated. Given the strong connections of the ANT with the hippocampus, its stimulation may have directly interfered with hippocampal function^90^ and contributed to symptom onset in Cases #17-22 together with MD stimulation. In some cases, symptoms might also have resulted from the disruption of one circuit in the presence of underlying abnormalities in the other. This may apply to the STN-DBS cases (Cases #25-29), in which a disruption of SNr output might have occurred in the context of a sensitized mesolimbic dopaminergic system.^91^ While the current data suggests that normalizing activity in the VTA-NAc loop (Cases #1,5) or modulating the SNr inhibitory output to the MD (Cases #12-14) may be effective in relieving symptoms^13,15–17^, further research should investigate whether combined modulation of both circuits could constitute a more powerful therapeutic strategy.

While we see great value in collating these cases given the key role of serendipitous observations in defining and refining DBS targets historically^23–26^, several limitations stem from the inherent nature of the data. First, our conclusions are based on the qualitative analysis of a small number of cases, given their rarity and despite extensive literature screening. Relevant cases related to other targets may not have been reported, potentially biasing anatomical inferences. Regardless, we see the main value of the present work in generating hypotheses about implicated circuits and potential surgical targets (with previous attempts remaining mostly theoretical^13,15^), which may be more directly tested in future work^19,92,93^ (Table S15). Moreover, electrode reconstruction and estimation of stimulation volumes may carry imprecision despite use of a state-of-the-art neuroimaging pipeline^27,94^, and the causal relationship between stimulation and symptoms cannot be fully ascertained despite strict inclusion criteria.^40^ It also remains unclear why stimulation of these same regions can sometimes occur without eliciting similar side effects^b^, and conversely, why these side effects were sometimes—albeit rarely—observed with active contacts placed within seemingly unrelated circuits (e.g., motor circuits; Case #25). Additionally, the observation that modulation of the same circuit can both improve or cause symptoms may appear puzzling, but is consistent with previous observations, and may generally relate to baseline levels of circuit (dys)function.^95^ Finally, beyond positive symptoms, negative symptoms should be considered when designing new therapeutic interventions for treatment-resistant schizophrenia.^13^

In conclusion, we gathered the largest collection to date of cases in which psychotic symptoms were affected by DBS, either intentionally or incidentally, and characterized the stimulated anatomical structures. Based on these findings, we propose a preliminary theoretical framework linking two candidate circuits to the emergence and improvement of psychotic symptoms, thereby generating testable hypotheses that may guide future mechanistic and clinical studies. Our study also highlights the value in reporting serendipitous stimulation effects (and potentially in collating these in a multicenter registry), preferably with precise information on stimulation location.

## Data Availability

Individual participant data underlying this study, including relevant clinical and imaging data, are not publicly available due to patient privacy concerns and institutional restrictions. Data may be made available to qualified investigators upon reasonable request and subject to data use agreements with the corresponding institutions contributing each case.

## Acknowledgements

The authors wish to acknowledge all researchers who were contacted via e-mail and kindly helped confirm the unsuitability of non-included cases.

## Contributors statement

G.M.M., A.R.P., A.H. and S.H.S. conceived and designed the study. G.M.M. and A.R.P. coordinated data collection and performed data analysis. E.G.S., I.A.S. and K.B. assisted data analysis. A.R., I.A.-B., F.L.W.V.J.S., C.N., S.J., H.K., S.H.M.J.P., L.M.R., M.J.K., L.L.G., P.E.M., A.R.C., K.O.D., J.B., A.S.W., D.D.D., Y.T., R.P.W.R., A.C., A.K., M.M., A.M., J.O.F., G.O., N.H., K.L., J.P., H.A., T.F., C.Z., J.A.A.-D., and I.C. contributed to case identification, data collection and verification at participating centers. G.M.M. and A.R.P. drafted the first version of the manuscript. All authors contributed to data interpretation, critically revised the manuscript for important intellectual content, and approved the final version for submission. G.M.M. and A.R.P. had full access and verified all data in the study.

## Footnotes

a For this reason, and because acetylcholine release is unlikely to be the main mechanism of DBS, especially in the context of early cholinergic degeneration in PD, stimulation of the NBM itself was not hypothesized to be the primary mechanism underlying hallucination relief in these two cases.

b We note that this seems to be the norm rather than the exception for DBS side effects; for example, mania has been linked to limbic STN stimulation, but not all patients with stimulation impinging on the limbic STN develop mania or hypomania.^50^

