## Supplementary material for "Deep brain stimulation and psychosis: A case series and two candidate causal brain circuits"

### Supplementary Material
#### Deep brain stimulation identifies two candidate circuits involved in psychosis

Garance M. Meyer*, Andrew R. Pines*, et al.

**Contents**

**Supplementary Methods**
Literature Search
Electrode reconstruction and calculation of stimulation volumes
**Table S1:** Summary of a-priori neurobiological models on schizophrenia and psychosis
**Figure S1:** Flow chart of the literature search procedure.
**Table S2:** Summary of cases receiving DBS for the treatment of psychosis.
**Table S3:** Summary of incidental cases.
**Figure S2** and **Table S4:** Stimulation volume overlaps for Cases #1-7 (NAc- and sgACC-DBS)
**Table S5:** White matter engagement for Cases #1-7 (NAc- and sgACC-DBS)
**Figure S3:** Stimulation volume overlaps for Cases #12-14 (SNr-DBS)
**Figure S4** and **Table S6:** Stimulation volume overlaps for Cases #15-16 (DBS of the habenula)
**Figure S5** and **Table S7:** Stimulation volume overlaps for Cases #17-22 (ANT-DBS)
**Table S8:** White matter engagement for Cases #17-22 (ANT-DBS)
**Figure S6:** Position of a typical CM-DBS electrode within the thalamus.
**Figure S7** and **Table S9:** Stimulation volume overlaps for Cases #25-29 (STN-DBS)
**Table S10:** White matter engagement for Cases #25-29 (STN-DBS)
**Figure S8** and **Table S11:** Stimulation volume overlaps for Case #31 (NAc/ALIC-DBS)
**Figure S9** and **Table S12:** Stimulation volume overlaps for Case #33 (dislodged GPi electrode)
**Figure S10** and **Table S13:** Stimulation volume overlaps for Case #34 (GPi-DBS)
**Figure S11** and **Table S14:** Stimulation volume overlaps for Case #35-36 (NBM-DBS)
**Supplementary material 2:** Early attempts at deep brain stimulation for psychosis
**Figure S12:** Approximative location of stimulation sites in Heath's first series of patients.
**Table S15**: Main hypotheses derived from the present study, and proposed follow-up studies.
**Supplementary references**

### Supplementary material S1: Supplementary Methods

#### Literature Search

We searched the PubMed database with the terms “DBS or deep brain stimulation” (Title/Abstract) and “psychosis or psychotic or hallucinations or delusions or schizophrenic or schizophrenia” (Title/Abstract). The search was first performed on November 11th, 2022, and updated on May 9th, 2024, for a total of 332 results.

Inclusion criteria were: 1) patient(s) with DBS irrespective of surgical target and indication or primary diagnosis, presenting with psychotic symptoms after DBS implantation (ON stimulation), with plausible causal relationship with the stimulation itself; or 2) patient(s) with DBS irrespective of surgical target and indication or diagnosis, presenting with improvement of their pre-existing psychotic symptoms after DBS implantation (ON stimulation), with plausible causal relationship with the stimulation itself; or 3) patients receiving DBS for treatment-resistant schizophrenia or psychosis. Exclusion criteria were: language other than English, animal research study, conference abstract, review article, duplicate report of same patient(s). Cases were included if psychotic symptoms of hallucinations, delusions, or both were present. This was based on previous findings that lesions causing different symptoms of psychosis map to common functional connections (Pines et al., 2025). Consistent with this previous work, thought disorder was present in some cases, but we did not include any case in which this was the only symptom, as it might be more difficult to operationalize and reliably identify, especially in the context of DBS side-effects. Causality was deemed plausible if symptoms started or worsened shortly after initiating DBS or shortly after a change in stimulation settings, and/or improved with further changes in settings or discontinuation. The same rationale was used for cases where DBS putatively improved psychotic symptoms (N=2, see below). Cases for which no sufficient elements to assess causality were provided, or for which changes in symptoms was most likely unrelated to DBS were excluded. Potential sources of bias and confounds were considered, such as concomitant changes in medication, potential metabolic and infectious causes of psychosis, and natural course of the primary disease. For example, we excluded cases of patients with Parkinson’s Disease who developed psychosis long after the surgery, with no mention of changes in stimulation parameters, which may be attributed to the natural course of the disease (Ffytche et al., 2017; Pagonabarraga et al., 2024), or cases of patients with epilepsy in which there were no clear arguments for a causal relationship with the stimulation itself (Nadkarni et al., 2007). Assessment of causality was performed by two raters (A.R.P., G.M.M.) before a consensus was reached. While this assessment may have eliminated potentially relevant cases, our priority was to focus on cases where stimulation was the most likely cause of the change in symptoms.

Given the rarity of these cases, the reference lists of the included publications as well as of reviews discussing side effects of DBS were screened for additional cases, and additional PubMed searches with terms like “psychiatric complications” or “adverse effects” were performed. Additionally, two unpublished cases were obtained from collaborators.

Authors of suitable reports were contacted via e-mail by the team at Brigham and Women's Hospital and offered to take part in the project by providing information regarding electrode position. In some cases, when causality appeared possible, but details were lacking to fully assess it, authors were contacted for more information. They were later offered to take part in the project if the report was deemed suitable. A flow chart is available as Fig. S1.

**Electrode reconstruction and calculation of stimulation volumes**

Electrode reconstruction was performed using the Lead-DBS toolbox (v3; www.lead-dbs.org; Neudorfer et al., 2023) based on the preoperative and postoperative imaging. Depending on data sharing restrictions, this was performed either by researchers at the corresponding institutions (who then provided the resulting MNI coordinates of the electrodes), or by the Brigham and Women's Hospital team.

The standard analysis pipeline was used. Briefly, preoperative and postoperative images were co-registered (using SPM12 for MRI scans, Friston et al., 2007, and ANTs for CT scans, Avants et al., 2011) and normalized to template "MNI" space (ICBM 2009b Nonlinear Asymmetric, ANTs-SyN algorithm, Avants et al., 2011, with default Lead-DBS presets, Ewert et al., 2019). Non-linear brain shift correction was applied on the postoperative CT scans to account for potential pneumocephalus (Horn & Kühn, 2015). Finally, the PaCER (Husch et al., 2018, for CT scans) or TRAC/CORE (Horn & Kühn, 2015), for MRI scans) algorithm was used to reconstruct the DBS electrodes, and when applicable, electrode orientation was detected using the DiODe algorithm (Dembek et al., 2021). Image quality and results of each step were carefully verified by visual inspection. Manual refinements of normalization and electrode reconstruction were performed as needed using dedicated tools (Neudorfer et al., 2023; Oxenford et al., 2024).

In some cases, approximative electrode reconstructions were obtained based on imaging or electrode coordinates available in the respective publications. For cases #8, 12, 13, 14 and 18, approximate MNI coordinates of the electrode contacts were determined based on the images of the MRI scans available in the original reports (with at least two different planes being shown) (Bioque et al., 2023; Doležalová et al., 2019; M. J. Kim et al., 2025).

The stimulation volumes were modeled using the OSS-DBS software (Butenko et al., 2020). Briefly, the method employs a finite element model to estimate the E-field in a 3D domain represented by a curved mesh with discrete conductivity values defined for cerebrospinal fluid, white matter, and grey matter. The calculations were performed in the native patient space when possible, and the resulting E-field magnitude maps were warped to MNI space using the previously computed transformation matrices. For the patients who received DBS for schizophrenia, clinical parameters were used. For the incidental cases, this was based on stimulation parameters putatively causing (or improving) the symptoms. When available, stimulation volumes were also modeled based on stimulation parameters not causing (or improving) the symptoms, for comparison.

| Neurobiological hypotheses | Relevant brain structures or pathways | Atlas resource |
|---|---|---|
| Dopamine hypothesis of schizophrenia - Hyperdopaminergic state in the mesolimbic system (Davis et al., 1991; Howes & Kapur, 2009)<br>• Effect of antipsychotics (D2r antagonists) (Seeman & Lee, 1975; van Rossum, 1966)<br>• Higher striatal dopamine synthesis and release capacity in patients with schizophrenia (molecular imaging) (Howes et al., 2012; McCutcheon et al., 2018)<br>• Some of the gene variants most strongly associated with schizophrenia are directly implicated in DA transmission, including DRD2 (Ripke et al., 2014; Trubetskoy et al., 2022) | Nucleus accumbens<br>Ventral pallidum<br>Ventral tegmental area<br>Medial forebrain bundle (ventral tegmental area projection pathway) | CIT168 (Pauli et al., 2018)<br>CIT168 (Pauli et al., 2018)<br>CIT168 (Pauli et al., 2018)<br>FOCUS atlas; based on (Coenen et al., 2023, 2024; Mai et al., 2015; Nieuwenhuys et al., 2008; Skandalakis et al., 2024) |
| Dopaminergic abnormalities localized to the associative, rather than limbic striatum, in schizophrenia<br>• One meta-analysis showing more elevated presynaptic dopamine functioning in the associative than in the limbic striatum in patients with schizophrenia based on N=7 molecular imaging studies (McCutcheon et al., 2018) | Caudate nucleus and putamen | CIT168 (Pauli et al., 2018) |
| Hippocampus abnormalities (Knight et al., 2022; Tamminga et al., 2010) - Hippocampal dysregulation of dopaminergic transmission (Grace et al., 2007; Lodge & Grace, 2007, 2011)<br>• Excess metabolic activity in the hippocampus; hyperactive hippocampus (Schobel et al., 2013)<br>• Hippocampus volume reductions in patients with schizophrenia (imaging and post-mortem studies) (Adriano et al., 2012; Roeske et al., 2021; van Erp et al., 2016)<br>• Lesions causing psychosis are functionally connected to the posterior hippocampus (Pines et al., 2025)<br>• Hippocampal stimulation normalizes DA transmission and behavior in a rat model of schizophrenia (Perez et al., 2012) | Hippocampus<br>Fornix | Juelich atlas (Amunts et al., 2005)<br>FOCUS atlas; based on (Ferreira et al., 2020; Mai et al., 2015) |

| | | |
|---|---|---|
| Glutamate hypothesis of schizophrenia (Kantrowitz & Javitt, 2010)<br>• Altered glutamate levels in schizophrenia (Merritt et al., 2023)<br>• NMDA receptor antagonists inducing psychotic-like symptoms (Beck et al., 2020)<br>• Various changes in NMDA receptor expression (post-mortem studies) (Hu et al., 2015)<br>• Some of the genes variants associated with schizophrenia are implicated in glutamatergic transmission (Ripke et al., 2014; Trubetskoy et al., 2022)<br>• NMDA receptor dysfunction in the hippocampus in schizophrenia (Beck et al., 2021; Lieberman et al., 2018; Pilowsky et al., 2006) | Poor localizing value as glutamate is the most common excitatory neurotransmitter in the brain, although the hippocampus may be especially sensitive to glutamatergic abnormalities (Lieberman et al., 2018; Tamminga et al., 2012) | |
| Thalamic dysfunction and abnormalities, centered on the MD and pulvinar (Alelú-Paz & Giménez-Amaya, 2008; Byne et al., 2009; Jiang et al., 2021; Pakkenberg et al., 2009; Steullet, 2020) - Thalamo-frontal cortical dysconnectivity and "hypofrontality" (Davis et al., 1991; Pakkenberg et al., 2009)<br>• Lesions of the MD causing psychotic symptoms (S. Kim & Kim, 2015; Kumral, 2001; Kumral & Oztürk, 2004; Liao et al., 2018; Little et al., 1986; Mäkelä et al., 1998; Malamud, 1967; Pavesi et al., 2014)<br>• Lesions causing delusions are functionally connected to the MD (Pines et al., 2025)<br>• Volume reductions of the MD and pulvinar in patients with schizophrenia, post-mortem and imaging evidence (Cullen et al., 2003; Dönmezler et al., 2024; Dorph-Petersen & Lewis, 2017; Gilbert et al., 2001; Mørch-Johnsen et al., 2023; Perez-Rando et al., 2022; Pergola et al., 2015, 2017; van Erp et al., 2016; Young et al., 2000)<br>• Abnormal metabolism in the MD in schizophrenia (Hazlett et al., 2004)<br>• Alterations of thalamo-frontal cortical functional connectivity (dMRI and fMRI) (Ellison-Wright & Bullmore, 2009; Pergola et al., 2015; Ramsay et al., 2023; Welsh et al., 2010; Woodward & Heckers, 2016)<br>• Improvement of pre-pulse inhibition by MD stimulation in a rat model of schizophrenia (Klein et al., 2013) | Mediodorsal thalamus, magnocellular and parvocellular parts;<br>Pulvinar<br><br>Substantia nigra pars reticulata<br>Nigrothalamic projections to mediodorsal thalamus<br><br>Amygdala<br>Ventral amygdalofugal pathway | Morel atlas (Krauth et al., 2010)<br><br>CIT168 (Pauli et al., 2018)<br>FOCUS atlas; based on (Carpenter et al., 1976; Carpenter & Peter, 1972; Erickson et al., 2004; François et al., 2002; Haber & Fudge, 1997; Ilinsky et al., 1985)<br><br>Jülich Atlas (Amunts et al., 2005)<br>FOCUS atlas; based on (Aggleton & Mishkin, 1984; Erkan et al., 2024; Klingler & Gloor, 1960; Li et al., 2020; Mai et al., 2015) |

| | | |
|---|---|---|
| Role of cholinergic transmission in visual hallucinations in PD (Barrett et al., 2018; Ffytche et al., 2017; Ignatavicius et al., 2025; Pagonabarraga et al., 2024) | Nucleus basalis of Meynert | Atlas of the human hypothalamus (Neudorfer et al., 2020) |
| Limbic dysregulation after STN-DBS in PD (Chopra et al., 2012; Prange et al., 2022) | Subthalamic nucleus, limbic territory<br>Limbic portion of the hyperdirect pathway<br>Medial forebrain bundle (ventral tegmental area projection pathway) | DISTAL atlas (including sub-territories; (Ewert et al., 2018)<br>FOCUS atlas (Rajamani et al., 2024)<br>FOCUS atlas; based on (Coenen et al., 2023, 2024; Mai et al., 2015; Nieuwenhuys et al., 2008; Skandalakis et al., 2024) |

**Table S1:** Summary of a-priori neurobiological models of schizophrenia and psychosis, non-exhaustive summary of evidence supporting each model, corresponding anatomical structures and pathways of interest, and atlases and normative connectome resources used for analyses.

| Case # Study | Target Indication Sex, Age | Case summary | Electrode Available imaging | Parameters | Right parameters | Left parameters |
|---|---|---|---|---|---|---|
| **#1** Corripio et al., 2020 (N1) | NAc SCZ F, 40-49 | There was a 40.2% improvement on the total PANSS (97 → 58) and 55.6% improvement in positive symptoms (27 → 12) during the stabilization phase. A worsening of symptoms was seen within 6 weeks when entering the crossover phase. | MedT 3387 T1, T2w MRI – CT | Clinical | C+ 8,9-, 2.5 V 150Hz, 130μs | C+ 1,2,3-, 2.5 V 150Hz, 130μs |
| **#2** Corripio et al., 2020 (N2) | sgACC SCZ M, 30-39 | There was a 19.4% improvement on the total PANSS (108 → 87) and 34.6% improvement in positive symptoms (26 → 17) during the stabilization phase. A worsening of symptoms was seen within a few days when entering the crossover phase. | MedT 3387 T1, T2w MRI – CT | Clinical | C+ 8,9,10,11-, 5V 150Hz, 150μs | C+ 0,1,2,3-, 5 V 150Hz, 150μs |
| **#3** Corripio et al., 2020 (N4) | sgACC SCZ F, 50-59 | There was a 36.9% improvement on the total PANSS (84 → 53) and 31.8% improvement in positive symptoms (22 → 15) during the stabilization phase. A worsening of symptoms was seen within 2 weeks when entering the crossover phase. | MedT 3387 T1, T2w MRI – CT | Clinical | C+ 9,10,11-, 5.5V 180Hz, 210μs | C+ 1,2,3-, 5.5 V 180Hz, 210μs |
| **#4** Corripio et al., 2020 (N5) | NAc SCZ M, 40-49 | There was a 22.5% improvement on the total PANSS (102 → 79) and 25.0% improvement in positive symptoms (24 → 18) during the stabilization phase. Incidental deactivation of the device had no obvious effect on symptoms. | MedT 3387 T1, T2w MRI – CT | Clinical | C+ 8,11-, 4 V 210Hz, 210μs | C+ 0,2-, 4 V 210Hz, 210μs |
| **#5** Corripio et al., 2020 (N6) | NAc SCZ F, 30-39 | There was a 49.2% improvement on the total PANSS (65 → 33) and 58.8% improvement in positive symptoms (17 → 7) during the stabilization phase. Symptoms returned within 24h when the device was accidentally switched off unbeknownst to the patients and investigators, and improved within 24h of restarting the stimulation. The patient did not enter the crossover phase. | MedT 3387 T1, T2w MRI – CT | Clinical | C+ 8,10-, 3.5 V 180Hz, 120μs | C+ 0,2-, 3.5 V 180Hz, 120μs |
| **#6** Corripio et al., 2020 (N7) | sgACC SCZ F, 30-39 | There was a 5.9% improvement on the total PANSS (85 → 80) and 14.8% improvement in positive symptoms (27 → 23) during the stabilization phase. The patient did not enter the crossover phase. | MedT 3387 T1, T2w MRI – CT | Clinical | C+ 9,10-, 4.5 V 210Hz, 180μs | C+ 1,3-, 4.5 V 210Hz, 180μs |

| | | | | | | |
|---|---|---|---|---|---|---|
| **#7**<br>Corripio et al., 2020<br>(N8) | sgACC<br>SCZ<br>M, 50-59 | There was a 16.1% improvement on the total PANSS (87 → 73) and 18.2% improvement in positive symptoms (22 → 18) during the stabilization phase. The patient did not enter the crossover phase. | MedT 3387<br>T1, T2w<br>MRI – CT | Clinical | C+ 9,11-, 4.5 V<br>180Hz, 210µs | C+ 1,3-, 4.5 V<br>180Hz, 210µs |
| **#8**<br>Bioque et al., 2023<br>(N2) | NAc<br>SCZ<br>M, 50-59 | The patient had treatment resistant schizophrenia. At 12 mo, there was a 29% improvement on the PANSS (140 → 100). | MedT 3389<br>NA | Clinical | NA, 4.5 V<br>130Hz, 90µs | NA, 4.5 V<br>130Hz, 90µs |
| #9<br>Bioque et al., 2023<br>(N3) | NAc<br>SAD<br>F, 40-49 | The patient had treatment resistant schizoaffective disorder. At 12 mo, there was a 13% improvement on the PANSS (91 → 79). | MedT 3389<br>NA | Clinical | NA, 3.5 V<br>130Hz, 90µs | NA, 3.5 V<br>130Hz, 90µs |
| #10<br>Bioque et al., 2023<br>(N4) | NAc<br>SCZ<br>M, 40-49 | The patient had treatment resistant schizophrenia. At 12 mo, there was a 15% improvement on the PANSS (107 → 91). | MedT 3389<br>NA | Clinical | NA, 7 V<br>130Hz, 90µs | NA, 7 V<br>130Hz, 90µs |
| #11<br>Zhou et al., 2020 | NAc/ALIC<br>AVH<br>(TBI)<br>F, 70-79 | The patient had auditory hallucinations, delusions (strange voices, thought broadcasting), depression and anxiety consecutive to traumatic brain injury. All symptoms were significantly improved at 18-mo follow-up (64.5% improvement on the PANSS; 96 → 34). | PINS L302<br>NA | Clinical<br>(18 mo) | "Point 3", 2.5 V<br>160Hz, 240µs | "Point 3", 2.5 V<br>160Hz, 240µs |
| **#12**<br>Cascella et al., 2021;<br>Kim et al. 2025 | SNr<br>SCZ<br>F, 30-39 | The patient had treatment resistant schizophrenia (auditory and visual hallucinations, thought broadcasting, persecutory delusions). DBS resulted in acute resolution of hallucinations, in a reproducible fashion. Hallucinations returned upon blinded temporary OFF stimulation. Positive symptoms were markedly improved at 3-mo, 6-mo and 1-y follow-up. At 6-mo, BPRS ratings for unusual thought content, hallucinations and delusional suspiciousness were improved by 84%. | MedT 3387<br>NA | Clinical | C+9-, 0.8 V<br>130 Hz, 60 µs | C+1-, 1 V<br>130 Hz, 60 µs |
| **#13**<br>Kim et al. 2025 | SNr<br>SCZ<br>F, NA | The patient had schizophrenia with auditory hallucinations. DBS resulted in acute reduction in the intensity of hallucinations during monopolar review. Long-term outcomes were not reported. | MedT 3387<br>NA | Monopolar review | NA | NA |

| | | | | | | |
|---|---|---|---|---|---|---|
| **#14**<br>Kim et al.<br>2025 | SNr<br>SCZ<br>M, NA | The patient had schizophrenia with auditory hallucinations. DBS resulted in acute reduction in the intensity of hallucinations during monopolar review. Long-term outcomes were not reported. | MedT 3387<br>NA | Monopolar review | NA | NA |
| **#15**<br>Wang et al., 2020<br>(Pt. 1) | Hb<br>SCZ<br>M, 20-29 | The patient had treatment resistant schizophrenia (auditory hallucinations, persecutory delusions, impulsive and abnormal behaviors, negative symptoms). DBS resulted in a modest improvement in positive symptoms at 7 mo, which was followed by marked worsening at 10 mo, leading to withdrawal from the study (9.5% worsening on the total PANSS, 74 → 81, and 69% worsening in positive symptoms, 13 → 22). | MedT 3389<br>T1, T2w<br>MRI - CT | Clinical | C+0-, 2.5 V<br>60 Hz, 60 µs | C+8-, 2 V<br>60 Hz, 60 µs |
| **#16**<br>Wang et al., 2020<br>(Pt. 2) | Hb<br>SCZ<br>M, 20-29 | The patient had treatment resistant schizophrenia (auditory hallucinations, persecutory delusions, destructive and aggressive behaviors, suicidal ideation). He demonstrated clinically significant improvement in positive and negative symptoms at 12 mo (31.7% improvement on the total PANSS, 120 → 82, 53.8% improvement in positive symptoms, 26 → 12). | PINS L301<br>T1, T2w<br>MRI - CT | Clinical | C+1,2-, 3.2 V<br>135 Hz, 60 µs | C+5,6-, 3.15 V<br>135 Hz, 80 µs |

**Table S2: Summary of cases receiving DBS for the treatment of psychosis.** Cases # in bold are the ones with available electrode reconstructions. The identifiers from the original publication are given in parentheses, when applicable. The table lists the intended DBS target as well as the primary indication for DBS (i.e., diagnosis). Age is given as range. Available imaging refers to the imaging data used for electrode reconstruction with the Lead-DBS software. Clinical stimulation parameters are given. Abbreviations: ALIC: Anterior limb of the internal capsule, AVH: Auditory verbal hallucinations, BPRS: Brief Psychiatric Rating Scale, F: Female, Hb: Habenula, M: Male, MedT: Medtronic, NAc: Nucleus accumbens, PANSS: Positive and Negative Syndrome Scale, SAD: Schizoaffective disorder, SCZ: Schizophrenia, sgACC: subgenual anterior cingulate (cg25), SNr: Substantia nigra pars reticulata, TBI: Traumatic brain injury.

| Case #<br>Study | Target<br>Indication<br>Sex, Age | Case summary | Electrode<br>Available imaging | Parameters | Right parameters | Left parameters |
|---|---|---|---|---|---|---|
| **#17**<br>Dague et al., 2023<br>(Pt. 7) | Epilepsy<br>ANT<br>M, 20-29 | The patient developed delusions and psychogenic non-epileptic seizures (PNES) approximately 20 months after DBS initiation. The symptoms persisted despite further changes in parameters. Delusions, but not PNES, disappeared when deactivating the device.<br>*History unspecified* | MedT 3387<br>T1, T2 and T2*w MRI - CT | Symptom onset | C+11-, 5 V<br>150 Hz, 90 µs | C+3-, 5 V<br>145 Hz, 90 µs |
| | | | | Resolution of symptoms | Deactivation of the device | |
| **#18**<br>Dolezalova et al. 2019 | Epilepsy<br>ANT<br>F, 40-49 | The patient gradually developed irritability, hostility and paranoia (delusions of control, e.g. DBS influencing her manners and actions and sending her thoughts to third party, persecutory delusions, e.g. someone spying on her) accompanied by thought disorder 3 months after implantation and 2 months after initiation of stimulation. Discontinuation of stimulation and antipsychotics only partially improved symptoms, with the patients requiring repeated psychiatric hospitalizations throughout the 7 years of follow-up.<br>*No history of psychosis* | MedT 3389<br>NA | Symptom onset | C+11-, 2.5 V<br>140 Hz, 90 µs | C+3-, 2.5 V<br>140 Hz, 90 µs |
| **#19**<br>Costa-Gertrudes et al., 2022<br>(Pt. 12) | Epilepsy<br>ANT<br>M, 20-29 | The patient developed psychosis that was relieved by reprogramming to more dorsal contact on the left electrode.<br>*History unspecified* | MedT 3389<br>T1, T2w, FGATIR MRI - CT | Symptom onset | C+2-, 5 V<br>140 Hz, 90 µs | C+10-, 5 V<br>140 Hz, 90 µs |
| | | | | Resolution of symptoms | C+2-, 5 V<br>140 Hz, 90 µs | C+11-, 5 V<br>140 Hz, 90 µs |
| **#20**<br>Järvenpää et al., 2018<br>(Pt. 1) | Epilepsy<br>ANT<br>F, 40-49 | The patient gradually developed psychotic symptoms in the form of delusions and erotomanic thoughts with anxiety, depression and irritability. Symptoms were relieved after DBS reprogramming to more dorsal contacts.<br>*No history of psychosis* | MedT 3389<br>T1w, STIR MRI - CT | Symptom onset | C+2-, 4 V / 4 mA<br>140 Hz, 90 µs | C+10-, 4 V / 6.6 mA<br>140 Hz, 90 µs |
| | | | | Resolution of symptoms | C+3-, 5.9 mA<br>140 Hz, 90 µs | C+11-, 6.3 mA<br>140 Hz, 90 µs |

| | | | | | | |
|---|---|---|---|---|---|---|
| **#21**<br>Järvenpää et al., 2018<br>(Pt. 3) | Epilepsy<br>ANT<br>M, 20-29 | The patient gradually developed psychotic symptoms in the form of delusions with disorganized thoughts, anxiety, low mood, issues with memory and irritability with aggressive thoughts. Symptoms were relieved after DBS reprogramming to more dorsal contacts.<br>*No history of psychosis* | MedT 3387<br>T1w, STIR<br>MRI - CT | Symptom onset | C+1,2-, 6.5 V / 10 mA<br>140 Hz, 90 µs | C+9,10-, 6.5 V / 10 mA<br>140 Hz, 90 µs |
| | | | | Resolution of symptoms | C+3-, 6.5 V<br>140 Hz, 90 µs | C+11-, 6.5 V<br>140 Hz, 90 µs |
| **#22**<br>Schaper et al., 2020<br>(Pt. 9) | Epilepsy<br>ANT<br>M, 30-39 | The patient developed psychotic symptoms, including delusions, thought disorder, and auditory hallucinations, a few days after an increase in stimulation voltage. Symptoms disappeared a few days after the voltage was reduced to its initial value.<br>*No history of psychosis* | MedT 3389<br>T1, T2w<br>MRI - CT | Symptom onset | C+2-, 5.5 V<br>135 Hz, 90 µs | C+9-, 5.5 V<br>135 Hz, 90 µs |
| | | | | Previous setting and resolution of symptoms | C+2-, 5 V<br>135 Hz, 90 µs | C+9-, 5 V<br>135 Hz, 90 µs |
| #23<br>Voges et al., 2015<br>(Pt. 3) | Epilepsy<br>ANT<br>F, 40-49 | The patient developed depression, anxiety, and paranoia, which disappeared upon nocturnal reduction of voltage.<br>*History unspecified* | MedT?<br>NA | Symptom onset | unspecified, 5 V<br>145 Hz, 90 µs | unspecified, 5 V<br>145 Hz, 90 µs |
| | | | | Resolution of symptoms | 1 V at night<br>145 Hz, 90 µs | 1 V at night<br>145 Hz, 90 µs |
| #24<br>Andrade et al., 2006<br>(CM1) | Epilepsy<br>CM<br>M, 40-49 | The patient developed psychosis with auditory hallucinations (hearing messages through songs and TV, followed orders of commanding voice) after a change in stimulation frequency from 65 to 185 Hz. Stimulation was stopped for two weeks before being restarted at 65 Hz.<br>*History unspecified* | MedT 3387<br>NA | Symptom onset | 1-2+, 2.5 V<br>185 Hz, 90 µs | 1-2+, 2.5 V<br>185 Hz, 90 µs |
| | | | | Previous and following settings | 1-2+, 2.5 V<br>65 Hz, 90 µs | 1-2+, 2.5 V<br>65 Hz, 90 µs |
| **#25**<br>Ito et al., 2020<br>(Pt. 5) | PD<br>STN<br>M, 50-59 | The patient developed delusions and hallucinations within a day after initiating stimulation, which were alleviated within a week after stopping stimulation, and reoccurred upon re-initiation. Symptoms disappeared when switching to bipolar settings.<br>*No history of psychosis* | BSci Vercise Directed T1, T2w MRI - CT | Symptom onset | C+2,3,4-, 0.3 mA<br>NA | C+2,3,4-, 0.3 mA<br>NA |
| | | | | Resolution of symptoms | Bipolar, unspecified | |

| | | | | | | |
|---|---|---|---|---|---|---|
| **#26**<br>Macerollo et al., 2020 | PD<br>STN<br>F, 40-49 | The patient developed severe frightening visual and sensory/tactile hallucinations (characters and animals) and psychosis seven days after initiation of bilateral STN-DBS (40 days post-implantation). Switching DBS off and reducing dopaminergic medication was found to have little effect. Hallucinations improved with antipsychotic medication.<br>*No history of psychosis* | BSci Vercise Directed<br>T1, T2w MRI - CT | Symptom onset | C+8-, 0.5 mA<br>149Hz, 60 µs | 13,14,15+<br>10,11,12-, 0.5 mA<br>149Hz, 60 µs |
| **#27**<br>Mosley et al., 2018 | PD<br>STN<br>M, 60-69 | The patient developed psychotic symptoms of persecutory delusions (clinicians and wife conspiring with police, threatened to kill his wife) in the context of mania and impulsivity after activation of an additional, more ventral contact. Symptoms did not improve with prolonged cessation of stimulation and were managed with antipsychotics. The patient later underwent replacement of the right electrode and did not require further psychiatric care or ongoing medication.<br>*No history of psychosis* | MedT 3389<br>T1w, FLAIR MRI - CT | Symptom onset | C+10,11-, 2.4 V<br>130 Hz, 60 µs | C+1-, 1 V<br>130 Hz, 60 µs |
| | | | | Previous settings | C+11-, 2.4 V<br>130 Hz, 60 µs | C+1-, 1 V<br>130 Hz, 60 µs |
| **#28**<br>Neudorfer et al. 2019 (Pt. 1) | PD<br>STN/VIM<br>M, 50-59 | The patient developed psychosis in the context of mania and impulsivity upon initiation of stimulation. Symptoms subsided immediately when more proximal contacts were activated.<br>*No history of psychosis* | BSci Vercise T1, T2w MRI, CT - x-rays | Symptom onset | C+9-, 2 mA<br>174Hz, 90 µs | C+1-, 1.5 mA<br>174Hz, 90 µs |
| | | | | Resolution of symptoms | More dorsal active contacts | |
| **#29**<br>Goede et al. | PD<br>STN<br>M, 60-69 | The patient developed psychosis (persecutory delusions and paranoia, with visual hallucinations), impulsivity and irritability, shortly after an increase in stimulation amplitude. Symptoms resolved with reprogramming to more dorsal contacts.<br>*No history of psychosis* | MedT B33005<br>T1, T2w MRI - CT | Symptom onset | C+1a,b,c-, 3.3 V<br>180Hz, 60 µs | C+1a,b,c-, 3.7 V<br>180Hz, 60 µs |
| | | | | Resolution of symptoms | C+2a,b,c-, 2.6 mA<br>180Hz, 60 µs | C+2a,b,c- 2.8 mA<br>180Hz, 60 µs |

| | | | | | | |
|---|---|---|---|---|---|---|
| #30<br>Herzog et al., 2003 | PD<br>STN<br>F, 60-69 | The patient developed persecutory delusions (sons conspiring, trying to get her money by threat of force) and disorganized behavior and thoughts in the context of mania within 3 weeks after initiation of stimulation. Stimulation arrest was not tolerated for more than 30 min, and psychotic symptoms were treated with antipsychotics.<br>*No history of psychosis* | NA | Symptom onset | Unspecified monopolar, 2.6 V<br>130 Hz, 60 µs | Unspecified monopolar, 2.2 V<br>130 Hz, 60 µs |
| **#31**<br>Testo et al., 2020 | OCD<br>NAc/ALIC<br>M, 20-29 | The patient experienced acute onset of psychosis, characterized by delusional beliefs and behaviors (talking to trees, hugging trees to “get the evil out of him”) as well as thought disorder. Symptoms occurred approximately 18 months after implantation, shortly after an increase in stimulation amplitude. There were no signs of hypomania. Antipsychotic medication and discontinuation of stimulation led to resolution of the symptoms within a week. No return of psychotic symptoms was observed when re-activating DBS with change in active contacts two years later.<br>*No history of psychosis* | MedT 3387<br>T1, T2w<br>MRI - CT | Symptom onset | C+3-, 5 V<br>135 Hz, 90 µs | C+2-, 9 V<br>135 Hz, 90 µs |
| | | | | Previous settings | C+3-, 5 V<br>135 Hz, 90 µs | C+2-, 7 V<br>135 Hz, 90 µs |
| | | | | Two years later (no psychosis) | C+2-, 1 V<br>135 Hz, 150 µs | 3+0,1-, 1 V<br>135 Hz, 150 µs |
| #32<br>Graat et al., 2019 | MDD<br>NAc/ALIC<br>M, 30-39 | The patient was admitted to the psychiatric emergency service for acute onset of paranoid delusions (thinking that there was asbestos in his house, neighbors, physicians and police conspiring against him), thought disorders anxiety and agitation two years after surgery. Lowering stimulation current led to resolution of delusions within hours. Delusions re-occurred within days upon current increase aiming to control depressive symptoms. Current increase could finally be achieved with the addition of antipsychotic medication.<br>*No history of psychosis* | MedT 3389<br>NA | Symptom onset | C+0,1-, 7.5 V<br>130 Hz, 90 µs | C+0,1-, 7.5 V<br>130 Hz, 90 µs |
| | | | | Resolution of symptoms | C+0,1-, 3.5 V<br>130 Hz, 90 µs | C+0,1-, 3.5 V<br>130 Hz, 90 µs |

| | | | | | | |
|---|---|---|---|---|---|---|
| **#33**<br>Piacentini et al., 2008 | DYT<br>GPi → AMY<br>M, 30-39 | The patient developed marked symptoms of delusions (bizarre beliefs of having paranormal abilities and the power to heal people). He additionally had mood lability (abrupt episodes of rage and anxiety) and depression (suicidal ideation and spending most of his day in bed) Imaging showed displacement of the left electrode to the amygdala region. Symptoms were relieved by discontinuation of the stimulation and left electrode repositioning.<br>*No history of psychosis* | MedT 3389<br>T1, T2w<br>MRI - CT | Symptom onset<br><br>Before onset<br><br>Resolution of symptoms | C+2-, 2.4V<br>180 Hz, 150 µs<br><br>Same settings, before electrode displacement<br><br>Discontinuation of stimulation and electrode replacement | C+5-, 2.7 V<br>180 Hz, 180 µs |
| **#34**<br>Hanna et al., 2022 | PD<br>GPi<br>M, 60-69 | The patient experienced acute onset severe psychosis (paranoia, thoughts that his spouse would kill or harm him, people coming after him) with depressive symptoms and suicidality within a few hours of DBS initiation. Symptoms ceased abruptly when stimulation at the right lead was discontinued and returned immediately with re-initiation. Stimulation could be continued at the right electrode using more ventral contacts without recurrence of symptoms.<br>*No history of psychosis* | MedT 3387<br>T1, T2w, FLAIR MRI - MRI | Symptom onset<br><br>Symptom onset<br>Resolution of symptoms<br>Resolution of symptoms | C+2-, 2.2 V<br>130 Hz, 90 µs<br><br>C+2- or C+3-<br>C+0- or C+1-<br>C+1-, 1.5V<br>130Hz, 60 µs | C+2-, 2 V<br>130 Hz, 60 µs<br><br>OFF<br>OFF<br>C+ 2-, 1.5V<br>130Hz, 60 µs |
| **#35**<br>Gratwicke et al., 2018<br>(A) | PDD<br>NBM<br>NA | The patient experienced near-complete cessation of their pre-existing complex visual hallucinations under stimulation. Resurgence of hallucinations was observed when stimulation was temporarily turned off.<br>*VH at baseline* | MedT 3387<br>T1, PDw<br>MRI - MRI | Disappearance of VH<br><br>Reappearance of VH | C+8,9-, 1.5V<br>20 Hz, 60 µs<br><br>Temporary OFF | C+0,1-, 1.5 V<br>20 Hz, 60 µs |
| **#36**<br>Gratwicke et al., 2018<br>(D) | PDD<br>NBM<br>NA | The patient experienced near-complete cessation of their pre-existing complex visual hallucinations under stimulation. Resurgence of hallucinations was observed when stimulation was temporarily turned off.<br>*VH at baseline* | MedT 3389<br>T1, PDw<br>MRI - MRI | Disappearance of VH<br><br>Reappearance of VH | C+9-, 3 V<br>20 Hz, 60 µs<br><br>Temporary OFF | C+1-, 3 V<br>20 Hz, 60 µs |

**Table S3: Summary of incidental cases.** Cases # in bold are the ones with available electrode reconstructions. The identifiers from the original publication are given in parentheses, when applicable. Cases #17 to 34 developed psychosis under DBS, while for Cases #35 and 36, psychotic symptoms improved under DBS. The table lists the intended DBS target as well as the primary indication for DBS (i.e., diagnosis). Age is given as range. Unspecified history refers to cases where the absence of history of psychosis was not clearly stated, although such history was not mentioned as a potential explanation for the symptoms. Available imaging refers to the imaging data used for electrode reconstruction with the Lead-DBS software. Stimulation parameters corresponding to times of change in symptoms are given. Abbreviations: ALIC: Anterior limb of the internal capsule, AMY: Amygdala, ANT: Anterior nucleus of the thalamus, BSci: Boston Scientific, CM: Centromedian nucleus of the thalamus, DYT: Dystonia F: Female, GPi: Globus pallidus pars interna, M: Male, MDD: Major depressive disorder, MedT: Medtronic, NA: Not available / unspecified, NAc: Nucleus accumbens, NBM: Nucleus basalis of Meynert, OCD: Obsessive compulsive disorder, PD: Parkinson's disease, PDD: Parkinson's disease dementia, STN: Subthalamic nucleus, VH: Visual hallucinations, VIM: Ventral intermediate nucleus of the thalamus.

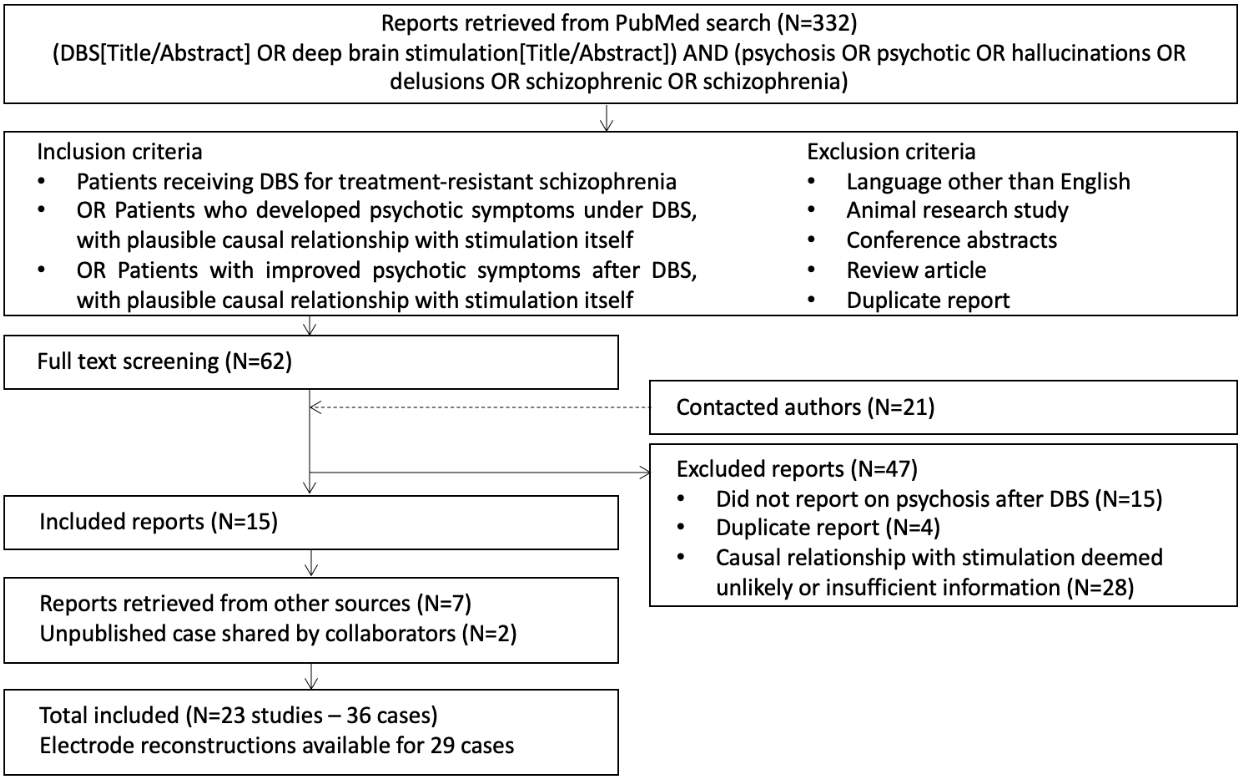


**Figure S1**: Flow chart of the literature search procedure.

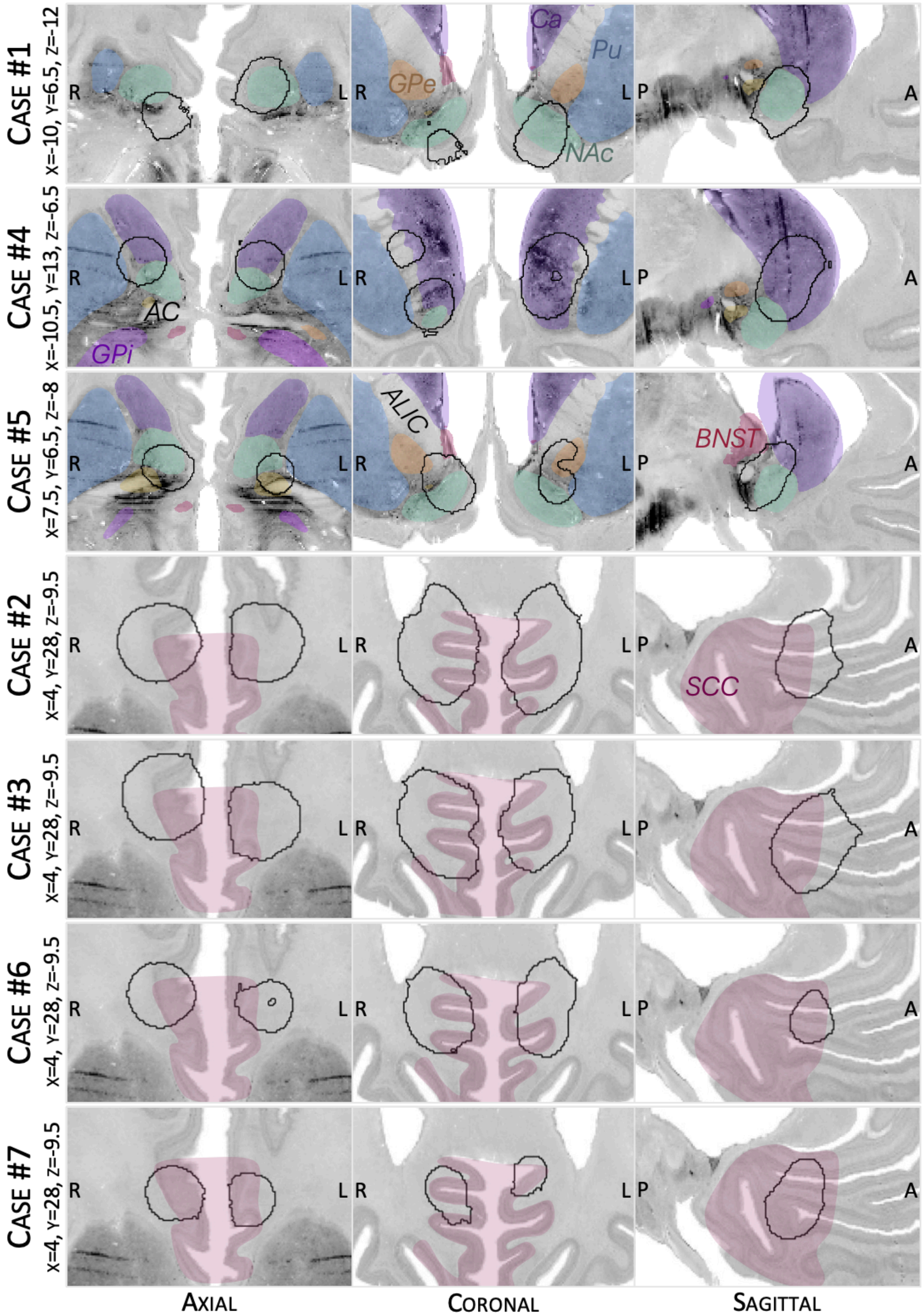


**Figure S2:** Overlap between stimulation volumes (black outlines) and surrounding anatomical structures for patients with DBS of the NAc and sgACC (Cases #1 to #7; see Table S2 for details). Structures are shown as defined in the DISTAL atlas, CIT168 atlas, and atlas of the human hypothalamus (Ewert et al., 2018; Neudorfer et al., 2020; Pauli et al., 2018). AC: anterior commissure, ALIC: anterior limb of the internal capsule, BNST: bed nucleus of the stria terminalis, Ca: caudate nucleus, GPe: globus pallidus pars externa, GPi: globus pallidus pars interna, NAc: nucleus accumbens, Pu: putamen, SCC: subcallosal cingulate.

| | NAc | Ca | Pu | VeP | GPe |
|---|---|---|---|---|---|
| Case #1 - NAc | | | | | |
| Right | 3.8% (18.2) | - | - | - | - |
| Left | 49.8% (346.2) | 8.0% (55.6) | 2.1% (14.8) | - | - |
| Case #4 - NAc | | | | | |
| Right | 14.2% (114.6) | 55.2% (447.3) | 3.7% (29.8) | - | - |
| Left | 6.8% (75.7) | 79.4% (884.5) | - | - | - |
| Case #5 - NAc | | | | | |
| Right | 27.9% (186.1) | 16.7% (111.8) | - | 2.8% (18.8) | 2.1% (14.0) |
| Left | 11.1% (54.3) | 12.9% (63.1) | 5.4% (26.2) | 7.7% (37.4) | 13.3% (65.1) |
| | BA24 | BA12 | BA33 | BA11 | BA32 |
| Case #2 - sgACC | | | | | |
| Right | 21.4% (470.1) | 12.0% (263.1) | 8.8% (193.0) | 5.2% (114.1) | - |
| Left | 27.0% (630.2) | 20.0% (467.0) | 13.4% (313.0) | 3.6% (84.3) | - |
| Case #3 - sgACC | | | | | |
| Right | 24.7% (530.3) | 19.7% (422.7) | 5.0% (108.0) | 13.6% (290.9) | 1.0% (22.5) |
| Left | 25.5% (494.3) | 14.0% (271.8) | 12.0% (233.0) | 19.1% (369.1) | - |
| Case #6 - sgACC | | | | | |
| Right | 30.9% (314.2) | 10.6% (107.5) | 4.8% (48.7) | 1.6% (15.8) | - |
| Left | 27.1% (264.8) | 11.3% (110.5) | 15.0% (146.1) | 1.0% (10.2) | - |
| Case #7 - sgACC | | | | | |
| Right | 30.7% (276.5) | 19.3% (174.1) | 6.2% (55.9) | 14.0% (126.6) | - |
| Left | 40.5% (301.9) | 6.5% (48.7) | 27.8% (207.4) | 2.1% (15.5) | - |

**Table S4:** Overlap between stimulation volumes ("VTAs") and surrounding anatomical structures for Cases #1 to #7 (NAc- and sgACC-DBS). Overlaps are given in percentage of total VTA volume and in $mm^3$ (in parentheses). Only structures with overlap of >1% of VTA are included. Anatomical structures were from the DISTAL atlas (Ewert et al., 2018), CIT168 atlas (Pauli et al., 2018), and atlas of the human hypothalamus (Neudorfer et al., 2020). BA: Brodmann area, Ca: caudate nucleus, GPe: globus pallidus pars externa, NAc: nucleus accumbens, Pu: putamen, VeP: ventral pallidum.

| | UF | Cingulum | Limbic HDP | Associative HDP | AC | MFB | Vafp | Fx |
|---|---|---|---|---|---|---|---|---|
| *NAc-DBS* | | | | | | | | |
| **Case #1** 40.2% improvement VTAs: 1181.1 $mm^3$ | | | 13.7% of fibers | | 3.7% of fibers | 44.8% of fibers | 17.5% of fibers | 3.1% of fibers |
| **Case #4** 22.5% improvement VTAs: 1934.5 $mm^3$ | | | 20.6 % of fibers | 25.7% of fibers | 1.8% of fibers | 75% of fibers | | |
| **Case #5** 49.2% improvement VTAs: 1152.0 $mm^3$ | | | 45.1% of fibers | 6.6% of fibers | 100% of fibers | 50% of fibers | 15.1% of fibers | 43.7% of fibers |
| *sgACC-DBS* | | | | | | | | |
| **Case #2** 19.4% improvement VTAs: 4527.2 $mm^3$ | 15.1% of fibers | 52% of fibers | 19.9% of fibers | | | | | |
| **Case #3** 36.9% improvement VTAs: 4089.8 $mm^3$ | 8.9% of fibers | 51% of fibers | 25.2% of fibers | | | | | |
| **Case #6** 5.9% improvement VTAs: 3165.3 $mm^3$ | 2.4% of fibers | 49% of fibers | 16.3% of fibers | | | | | |
| **Case #7** 16.1% improvement VTAs: 1650.0 $mm^3$ | | 46% of fibers | 24.4% of fibers | | | | | |

**Table S5:** Overlap between stimulation volumes ("VTAs") and surrounding white matter pathways for with DBS of the NAc and sgACC (Cases #1 to #7; see Table S2 for details). Only pathways with overlap of >1% of fibers are included. Results were pooled across hemispheres. VTA volume is given in $mm^3$ combined across hemispheres. AC: Anterior commissure, Cingulum: cingulum, para-olfactory segment, Fx: fornix, HDP: hyperdirect pathway (limbic: from BA 13, 24/32, 25; associative: from BA 10, 45/47), MFB: medial forebrain bundle – ventral tegmental area to nucleus accumbens, UF: uncinate fasciculus, Vafp: ventral amygdalofugal pathay.

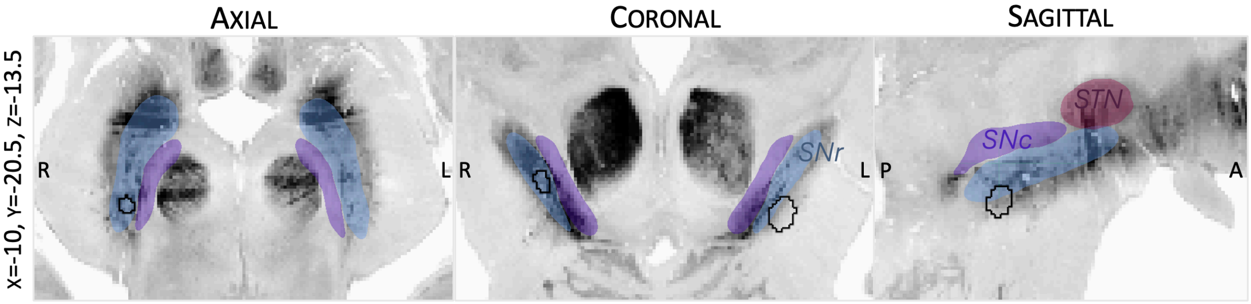


**Figure S3:** Overlap between stimulation volumes (black outlines) and surrounding anatomical structures for Case #12 (SNr-DBS, see Table S2). VTAs mainly overlapped with the SNr (96.9% and 16.0% of the right and left VTAs, respectively). Chronic stimulation parameters were not available for Cases # 13 and 14 such that VTAs could not be calculated. All electrodes had at least two contacts in the SNr. Structures are shown as defined in the DISTAL and CIT-168 atlases (Ewert et al., 2018; Pauli et al., 2018). SNc: substantia nigra pars compacta, SNr: substantia nigra pars reticulata, STN : subthalamic nucleus.

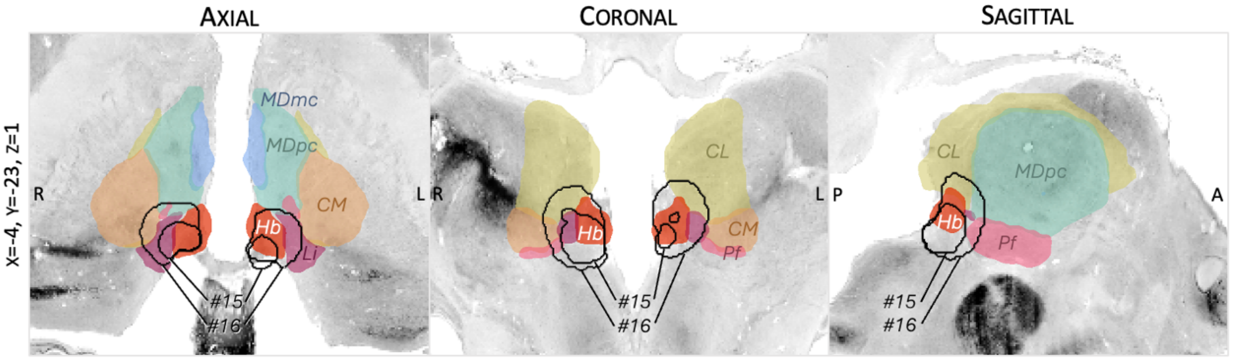


**Figure S4:** Overlap between stimulation volumes (black outlines) and surrounding anatomical structures for patients with DBS of the habenula (Cases #15, non-responder; and #16, responder, see Table S2). Structures are shown as defined in the Morel thalamic atlas (Jakab et al., 2012). CL: central lateral nucleus, CM: centromedian nucleus, Hb: habenula, Li: limitans nucleus, MDmc: magnocellular mediodorsal nucleus, MDpc: parvocellular mediodorsal nucleus.

| Right VTA | | Left VTA | |
|---|---|---|---|
| *Case #15* | | | |
| Hb | 36.0% - 14.4 $mm^3$ | Hb | 35.1% - 7.2 $mm^3$ |
| Li | 19.3% - 7.8 $mm^3$ | | |
| Pf | 1.4% - 0.6 $mm^3$ | | |
| *Case #16* | | | |
| CL | 23.4% - 34.7 $mm^3$ | Hb | 38.9% - 44.8 $mm^3$ |
| Hb | 22.6% - 33.5 $mm^3$ | CL | 25.6% - 29.5 $mm^3$ |
| Li | 18.9% - 28.0 $mm^3$ | Li | 17.8% - 20.5 $mm^3$ |
| Pf | 11.1% - 16.4 $mm^3$ | MDpc | 2.9% - 3.35 $mm^3$ |
| MDpc | 7.7% - 11.4 $mm^3$ | | |
| CM | 7.4% - 11.0 $mm^3$ | | |

**Table S6:** Overlap between stimulation volumes ("VTAs") and surrounding anatomical structures for patients with DBS of the habenula (Cases #15 and #16), in percentage of total VTA volume and in $mm^3$ (only structures with overlap of >1% of VTA are included). The Morel atlas was used (Jakab et al., 2012). CL: central lateral nucleus, CM: centromedian nucleus, Hb: habenula, Li: limitans nucleus, MDpc: parvocellular mediodorsal nucleus.

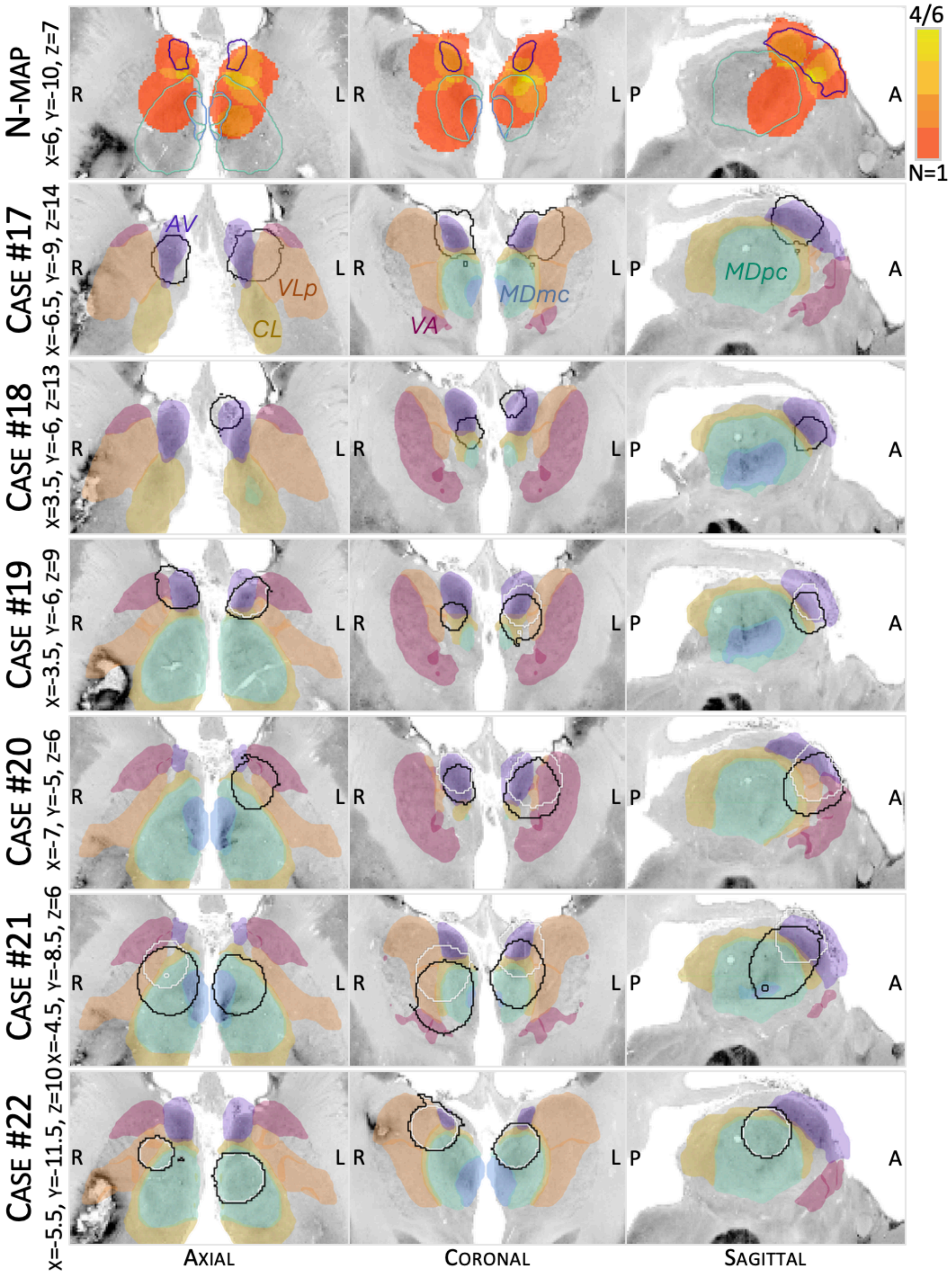


**Figure S5:** Overlap between stimulation volumes and surrounding anatomical structures for Cases #17-22 (ANT-DBS; see Table S3). Black outlines denote stimulation volumes associated with symptom onset, while white outlines denote stimulation volumes associated with symptom resolution. Structures are shown as defined in the Morel thalamic atlas (Jakab et al., 2012). AV: anterior ventral nucleus, CL: central lateral nucleus, MDmc: mediodorsal nucleus – magnocellular part, MDpc: mediodorsal nucleus – parvocellular part, VA: ventral anterior nucleus (magnocellular and parvocellular parts), VLp: ventral lateral posterior nucleus (dorsal and ventral parts).

| | AV | MDpc | MDmc | CL | VLpd | VLpv | VApc | VAmc | CeM | AM | AD | LD |
|---|---|---|---|---|---|---|---|---|---|---|---|---|
| **Case #17 - Symptom onset** | | | | | | | | | | | | |
| Right | 45.7% (108.6) | 5.3% - (12.5) | - | 15.7% (37.3) | 8.4% (19.9) | - | - | - | - | - | 2.4% (5.7) | 1.7% (4.1) |
| Left | 29.7% (99.2) | 2.6% (8.5) | - | 10.2% (34.2) | 50.3% (167.9) | - | 1.9% (6.5) | - | - | - | - | - |
| **Case #18 - Symptom onset** | | | | | | | | | | | | |
| Right | 65.3% (48.3) | 5.6% (4.2) | - | 22.8% (16.9) | - | - | - | - | 4.7% (3.5) | 8.3% (6.1) | - | - |
| Left | 52.4% (48.0) | - | - | - | - | - | - | - | - | - | - | - |
| **Case #19 - Symptom onset** | | | | | | | | | | | | |
| Right | 54.7% (124.4) | - | - | 7.4% (16.9) | 2.2% (4.9) | 4.0% (9.0) | 18.0% (40.9) | 4.4% (10.0) | - | 7.7% (17.6) | - | - |
| Left | 44.0% (97.2) | 6.1% (13.5) | - | 21.2% (46.8) | 4.4% (9.7) | 9.9% (21.9) | 12.8% (28.3) | 5.2% (11.6) | 2.7% (5.9) | 3.7% (8.2) | - | - |
| **Case #19 - Symptom resolution** | | | | | | | | | | | | |
| Right | 54.7% (124.4) | - | - | 7.4% (16.9) | 2.2% (4.9) | 4.0% (9.0) | 18.0% (40.9) | 4.4% (10.0) | - | 7.7% (17.6) | - | - |
| Left | 65.1% (115.6) | - | - | 10.6% (18.9) | 8.1% (14.4) | 5.7% (10.0) | 12.4% (21.9) | 1.6% (2.8) | - | - | - | - |
| **Case #20 - Symptom onset** | | | | | | | | | | | | |
| Right | 91.1% (105.8) | - | - | 7.9% (9.2) | - | - | 1.2% (1.4) | - | - | 1.9% (2.2) | - | - |
| Left | 24.1% (106.0) | 14.3% (63.0) | - | 17.6% (77.6) | 11.0% (48.3) | 16.5% (72.6) | 22.1% (97.6) | 2.8% (12.1) | - | - | - | - |

| | | | | | | | | | | | | |
|---|---|---|---|---|---|---|---|---|---|---|---|---|
| **Case #20 - Symptom resolution** | | | | | | | | | | | | |
| Right | 79.3% (109.8) | - | - | - | - | - | 2.7% (3.7) | - | - | - | - | - |
| Left | 32.1% (112.3) | 1.6% (5.6) | - | 9.5% (33.4) | 19.1% (66.8) | 11.8% (41.3) | 26.6% (93.1) | 1.0% (3.6) | - | - | - | - |
| **Case #21 - Symptom onset** | | | | | | | | | | | | |
| Right | - | 48.8% (397.0) | 13.5% (109.7) | 15.1% (122.8) | 1.7% (14.0) | 18.3% (149.1) | 4.0% (32.6) | 1.9% (15.1) | 7.0% (56.6) | - | - | - |
| Left | 14.2% (92.9) | 51.9% (338.4) | 15.5% (101.4) | 19.4% (126.5) | 4.3% (28.0) | 3.1% (20.3) | - | - | - | - | - | - |
| **Case #21 - Symptom resolution** | | | | | | | | | | | | |
| Right | 23.7% (112.2) | 22.8% (107.7) | - | 20.2 (95.5) | 14.4% (67.9) | 17.3% (81.8) | 10.4% (49.0) | 1.4% (6.5) | - | - | - | - |
| Left | 50.1% (170.9) | 8.7% (29.6) | - | 15.7% (53.6) | 15.5% (52.8) | 1.2% (3.9) | 1.5% (5.1) | - | - | - | 1.4% (4.7) | - |
| **Case #22 - Symptom onset** | | | | | | | | | | | | |
| Right | 21.1% (80.3) | 8.4% (32.1) | - | 14.5% (55.4) | 55.5% (211.3) | 2.0% (7.6) | - | - | - | - | - | - |
| Left | 2.3% (7.4) | 75.8% (242.8) | 4.2% (13.3) | 24.9% (80.0) | - | - | - | - | - | - | - | 1.1% (3.7) |
| **Case #22 - Previous settings and resolution of symptoms** | | | | | | | | | | | | |
| Right | 21.3% (69.7) | 7.6% (24.7) | - | 14.4% (47.0) | 58.0% (189.8) | 1.6% (5.4) | - | - | - | - | - | - |
| Left | 1.6% (4.7) | 77.7% (219.2) | 3.1% (8.8) | 24.6% (69.3) | - | - | - | - | - | - | - | 1.0% (2.9) |

**Table S7:** Overlap between stimulation volumes (“VTAs”) and surrounding anatomical structures for Cases #17 to #22 (ANT-DBS). Overlaps are given in percentage of total VTA volume and in $mm^3$ (in parentheses). Only structures with overlap of >1% of VTA are included. Anatomical structures were from the Morel thalamic atlas (Jakab et al., 2012). AD: anterior dorsal nucleus, AM: anterior medial nucleus, AV: anterior ventral nucleus, CeM: central medial nucleus, CL: central lateral nucleus, LD: lateral dorsal nucleus, MDmc: mediodorsal nucleus – magnocellular part, MDpc: mediodorsal nucleus – parvocellular part, VAmc: ventral anterior nucleus –

magnocellular part, VApc: ventral anterior nucleus – parvocellular part, VLpd: ventral lateral posterior nucleus – dorsal part, VLpv: ventral lateral posterior nucleus – ventral part.

| | Mmt | Nigrothalamic pathway to MD | Fx | Vafp | St |
|---|---|---|---|---|---|
| **Case #17 - Symptom onset** | 23.8% of fibers | | 4.2% of fibers | | 1.6% of fibers |
| **Case #18 - Symptom onset** | 45% of fibers | | 30.9% of fibers | | |
| **Case #19 - Symptom onset** | 100% of fibers | | | | |
| **Case #19 - Symptom resolution** | 100% of fibers | | | | |
| **Case #20 - Symptom onset** | 99.2% of fibers | 11.7% of fibers | | | |
| **Case #20 - Symptom resolution** | 97.4% of fibers | | | | 22.4% of fibers |
| **Case #21 - Symptom onset** | 75.4% of fibers | 97.1% of fibers | | 74.4% of fibers | |
| **Case #21 - Symptom resolution** | 93.6% of fibers | 18.8% of fibers | 6.5% of fibers | | |
| **Case #22 - Symptom onset** | 3.0% of fibers | | | 6.6% of fibers | |
| **Case #22 - Previous settings and resolution of symptoms** | 1.4% of fibers | | | 5.4% of fibers | |

**Table S8:** Overlap between stimulation volumes ("VTAs") and surrounding white matter pathways for Cases #17 to #22 (ANT-DBS). Only pathways with overlap of >1% of fibers are included. Results were pooled across hemispheres. Fx: fornix, Mmt: mammillothalamic tract, Nigrothalamic pathway: nigrothalamic pathway – substantia nigra pars reticulata to mediodorsal nucleus, St: stria terminalis, Vafp: ventral amygdalofugal pathay – amygdala to mediodorsal nucleus.

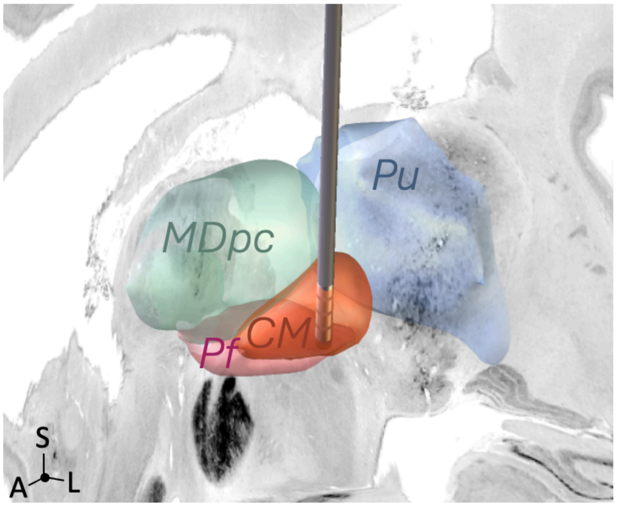


**Figure S6:** Position of a typical CM-DBS electrode within the thalamus. The typical CM-DBS target for epilepsy is 10 lateral and 0-1mm anterior to the posterior commissure (PC), at the level of PC for the tip of the electrode (Velasco et al., 2006; MNI coordinates: 10.5, -22, -2.1). Although electrode reconstruction was not available in Case #24, given that no other cases of psychosis were described after CM-DBS to the best of our knowledge (with a total of >200 cases of CM-DBS worldwide, (Hart et al., 2025), one may speculate that psychosis related to stimulation impinging on adjacent thalamic structures, such as the MD or pulvinar, which have been implicated in schizophrenia (Byne et al., 2009; Perez-Rando et al., 2022). Structures are shown as defined in the Morel thalamic atlas (Jakab et al., 2012). CM: centromedian nucleus, MDpc: parvocellular mediodorsal nucleus, Pf: Parafascicular nucleus, Pu: Pulvinar.

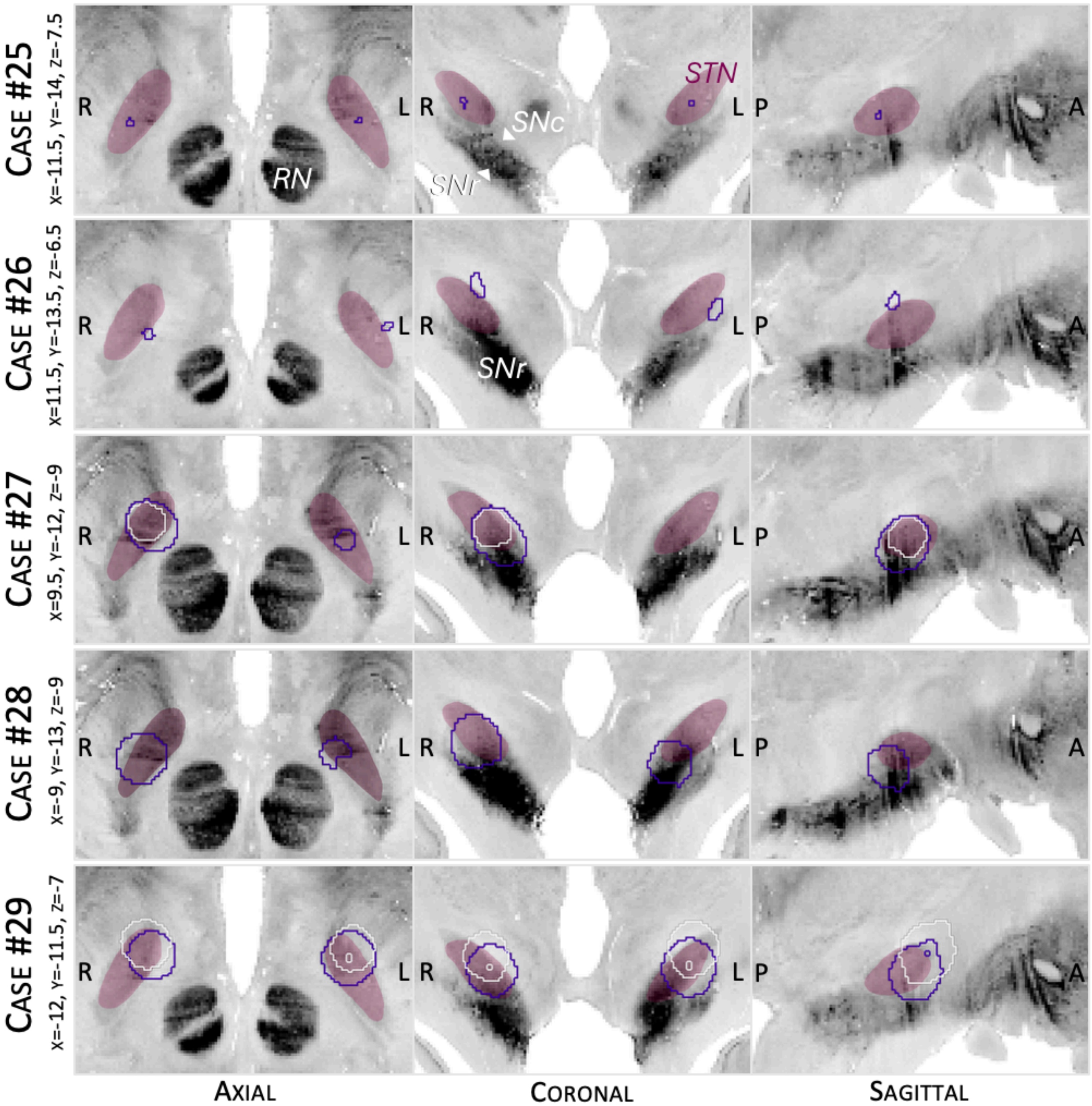


**Figure S7:** Overlap between stimulation volumes and surrounding anatomical structures for cases #25 to #29 (STN-DBS). Outlines represent stimulation volumes associated (purple) and not associated (white) with symptoms (see Table S3 for details). The subthalamic nucleus STN is shown from the DISTAL atlas (Ewert et al., 2018)

| | STNm | STNa | STNl | SNr | SNc |
|---|---|---|---|---|---|
| Case #25 - Symptom onset | | | | | |
| Right | 92.5% (1.2) | - | 7.5% (0.1) | - | - |
| Left | 89.0% (0.9) | 11.0% (0.1) | - | - | - |
| Case #26 - Symptom onset | | | | | |
| Right | 3.4% (0.2) | 12.7% (0.7) | - | - | - |
| Left | 2.0% (0.1) | - | 20.6% (1.3) | 24.0% (1.6) | - |
| Case #27 - Symptom onset | | | | | |
| Right | 3.6% (4.1) | 27.0% (30.3) | 24.7% (27.8) | 18.9% (21.2) | 2.7% (3.0) |
| Left | 56.0% (5.4) | 42.4% (4.1) | 1.4% (0.1) | 7.6% (0.7) | - |
| Case #27 - Previous settings | | | | | |
| Right | 2.7% (1.3) | 53.9% (26.4) | 35.1% (17.2) | 5.1% (2.5) | - |
| Left | 55.7% (5.4) | 42.8% (4.2) | 1.4% (0.1) | 7.4% (0.7) | - |
| Case #28 - Symptom onset | | | | | |
| Right | 33.3% (34.3) | 12.6% (13.0) | 16.8% (17.3) | 19.6% (20.2) | - |
| Left | 4.3% (2.4) | 14.9% (8.4) | 15.3% (8.7) | 39.9% (22.6) | 5.0% (2.8) |
| Case #29 - Symptom onset | | | | | |
| Right | 2.7% (2.5) | 40.1% (37.5) | 24.4% (22.8) | 1.1% (1.1) | - |
| Left | 14.1% (16.8) | 39.7% (47.5) | 5.0% (6.0) | 6.2% (7.5) | - |
| Case #29 - Symptom resolution | | | | | |
| Right | - | 35.3% (38.4) | 5.3% (5.7) | - | - |
| Left | 3.6% (4.4) | 22.2% (27.1) | 2.0% (2.4) | - | - |

**Table S9:** Overlap between stimulation volumes (“VTAs”) and surrounding anatomical structures for Cases #25 to #29 (STN-DBS). Overlaps are given in percentage of total VTA volume and in $mm^3$ (in parentheses). Only structures with overlap of >1% of VTA are included. Anatomical structures were from the DISTAL atlas (Ewert et al., 2018) and CIT168 atlas (Pauli et al., 2018). SNc: substantia nigra pars compacta, SNr: substantia nigra pars reticulata, STNa: subthalamic nucleus – associative territory, STNl: subthalamic nucleus – limbic territory, STNm: subthalamic nucleus – motor territory.

| | Nigrothalamic pathway to MD | Motor HDP | Associative HDP | Limbic HDP |
|---|---|---|---|---|
| Case #25 - Symptom onset | | | | |
| Case #26 - Symptom onset | | 1.3% of fibers | | |
| Case #27 - Symptom onset | 50% of fibers | 31.5% of fibers | 35.3% of fibers | 42.4% of fibers |
| Case #27 - Previous settings | 19.1% of fibers | 29.2% of fibers | 17.9% of fibers | 26.3% of fibers |
| Case #28 - Symptom onset | 56.4% of fibers | 26.1% of fibers | | 31.7% of fibers |
| Case #29 - Symptom onset | 48.4% of fibers | 66.0% of fibers | 34.5% of fibers | 40.3% of fibers |
| Case #29 - Symptom resolution | 1.5% of fibers | 66.4% of fibers | 30.5% of fibers | 29.8% of fibers |

**Table S10:** Overlap between stimulation volumes ("VTAs") and surrounding white matter pathways for Cases #25 to #29 (STN-DBS). Only pathways with overlap of >1% of fibers are included. Results were pooled across hemispheres. HDP: hyperdirect pathway (limbic: from BA 13, 24/32, 25; associative: from BA 10, 45/47; motor: from BA 4, 6, 8), Nigrothalamic pathway: nigrothalamic pathway – substantia nigra pars reticulata to mediodorsal nucleus.

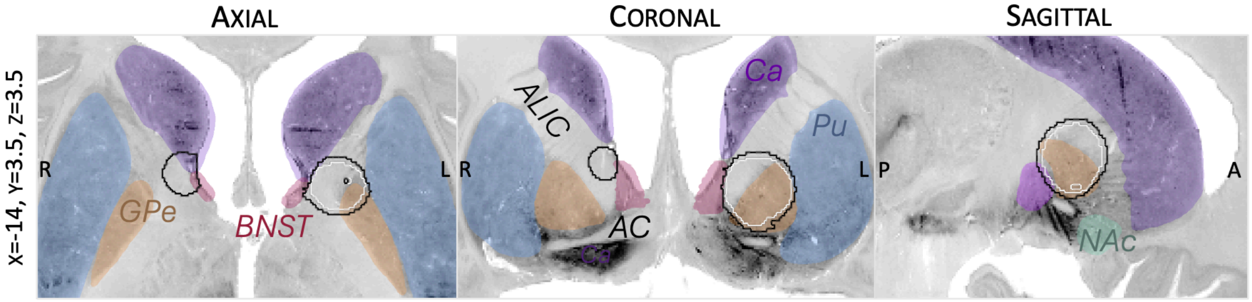


**Figure S8:** Overlap between stimulation volumes and surrounding anatomical structures for Case #31 (NAc/ALIC-DBS). Outlines represent stimulation volumes associated (black) and not associated (white, i.e. before change in parameters and symptom onset) with symptoms (see Table S3 for detail). Structures are shown as defined in the DISTAL atlas, CIT168 atlas, and atlas of the human hypothalamus (Ewert et al., 2018; Neudorfer et al., 2020; Pauli et al., 2018). AC: anterior commissure, ALIC: anterior limb of the internal capsule, BNST: bed nucleus of the stria terminalis, Ca: caudate nucleus, GPe: globus pallidus pars externa, GPi: globus pallidus pars interna, NAc: nucleus accumbens, Pu: putamen.

| Right VTA | | Left VTA | |
|---|---|---|---|
| *Symptom onset* | | | |
| Ca | 37.0% - 63.3 $mm^3$ | GPe | 46.3% - 373.7 $mm^3$ |
| BNST | 5.1% - 8.8 $mm^3$ | GPi | 1.9% - 15.6 $mm^3$ |
| | | Pu | 1.2 % - 10 $mm^3$ |
| *White matter pathways (bilateral)* | | | |
| St, 1.0% of fibers | | | |
| AC, 1.2% of fibers | | | |
| Motor HDP, 4.3% of fibers | | | |
| Associative HDP, 57.8% of fibers | | | |
| Limbic HDP, 30.4% of fibers | | | |
| *Previous settings* | | | |
| Ca | 37.0% - 63.3 $mm^3$ | GPe | 52.9% - 296.8 $mm^3$ |
| BNST | 5.1% - 8.8 $mm^3$ | | |
| *White matter pathways (bilateral)* | | | |
| Motor HDP, 1.3% of fibers | | | |
| Associative HDP, 55.6% of fibers | | | |
| Limbic HDP, 17.2% of fibers | | | |

**Table S11:** Overlap between stimulation volumes ("VTAs") and surrounding anatomical structures and white matter pathways for Case #31 (NAc-DBS). Structure overlaps are given in percentage of total VTA volume and in $mm^3$ (only structures with overlap of >1% of VTA and pathways with overlap of >1% of fibers are included). Anatomical structures were from the DISTAL atlas (Ewert et al., 2018), CIT168 atlas (Pauli et al., 2018), and atlas of the human hypothalamus (Neudorfer et al., 2020). AC : Anterior commissure, BNST: Bed nucleus of the stria terminalis, Ca: Caudate, GPe: Globus pallidus externus, GPi: Globus pallidus internus, HDP: hyperdirect pathway (limbic: from BA 13, 24/32, 25; associative: from BA 10, 45/47; motor: from BA 4, 6, 8), Pu: Putamen, St: Stria terminalis.

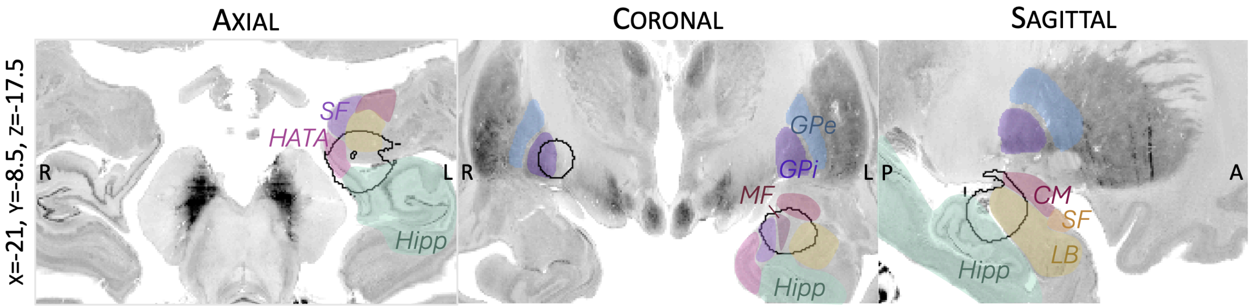


**Figure S9:** Overlap between stimulation volumes (black outlines) and surrounding anatomical structures for Case #33 (GPi electrode dislodged to amygdala region, see Table S3 for details). Structures are shown as defined in the DISTAL and Jülich atlases (Amunts et al., 2005; Ewert et al., 2018). CM: centromedian amygdala, GPe: Globus pallidus externus, GPi: Globus pallidus internus, HATA: Hippocampal-amygdaloid transition area, LB: Laterobasal amygdala, SF: Superficial amygdala, VTM: Ventromedial nucleus. Structures

| Right VTA | | Left VTA | |
|---|---|---|---|
| *Symptom onset* | | | |
| CA2 | 22.8% - 155.3 $mm^3$ | GPi | 55.6% - 82.9 $mm^3$ |
| HATA | 12.8% - 86.9 $mm^3$ | | |
| CM amygdala | 8.7% - 59.1 $mm^3$ | | |
| VTM amygdala | 4.3% - 29.5 $mm^3$ | | |
| MF amygdala | 3.1% - 21.2 $mm^3$ | | |
| SF amygdala | 2.8% - 18.9 $mm^3$ | | |
| CA1 | 2.5% - 16.9 $mm^3$ | | |
| LB amygdala | 2.0% - 13.3 $mm^3$ | | |

**Table S12:** Overlap between stimulation volumes ("VTAs") and surrounding anatomical structures for Case #33, in percentage of total VTA volume and in $mm^3$ (only structures with overlap of >1% of VTA are included). The Jülich and DISTAL atlases were used (Amunts et al., 2005; Ewert et al., 2018). CM: centromedian, GPe: Globus pallidus externus, GPi: Globus pallidus internus, HATA: Hippocampal-amygdaloid transition area, LB: Laterobasal, SF: Superficial, VTM: Ventromedial.

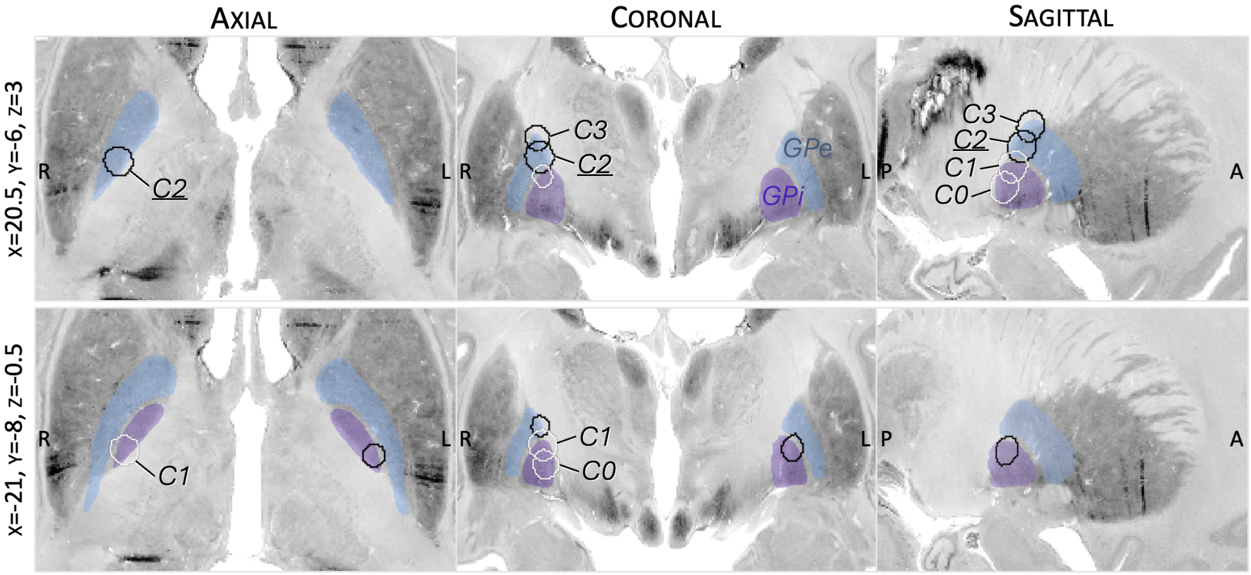


**Figure S10:** Overlap between stimulation volumes and surrounding anatomical structures for Case #34 (GPi-DBS). Outlines represent stimulation volumes associated (black) and not associated (white) with symptoms. Specifically, stimulation of contact C2 was associated with symptom onset, with the stimulation volume mostly involving the GPe. Attempts to stimulate contacts C2 or C3 led to reoccurrence of symptoms, while stimulation of contacts C0 or C1 did not (see Table S3 for detail). Structures are shown as defined in the DISTAL atlas (Ewert et al., 2018). GPe: Globus pallidus externus, GPi: Globus pallidus internus.

| Right VTA | | Left VTA | |
|---|---|---|---|
| *Symptom onset (C2)* | | | |
| GPe | 76.5% - 57.3 $mm^3$ | GPi | 76.7% - 31.5 $mm^3$ |
| GPi | 1.7% - 1.3 $mm^3$ | GPe | 6.4% - 2.6 $mm^3$ |
| *Symptom onset (C3)* | | | |
| GPe | 40.5% - 30.0 $mm^3$ | OFF | |
| *Resolution (C0)* | | | |
| GPi | 58.5% - 44.8 $mm^3$ | OFF | |
| *Resolution (C1)* | | | |
| GPi | 50.7% - 37.5 $mm^3$ | OFF | |
| GPe | 14.3% - 10.5 $mm^3$ | | |

**Table S13:** Overlap between stimulation volumes (“VTAs”) and surrounding anatomical structures for Case #34 (GPi-DBS), in percentage of total VTA volume and in $mm^3$. GPe: Globus pallidus externus, GPi: Globus pallidus internus.

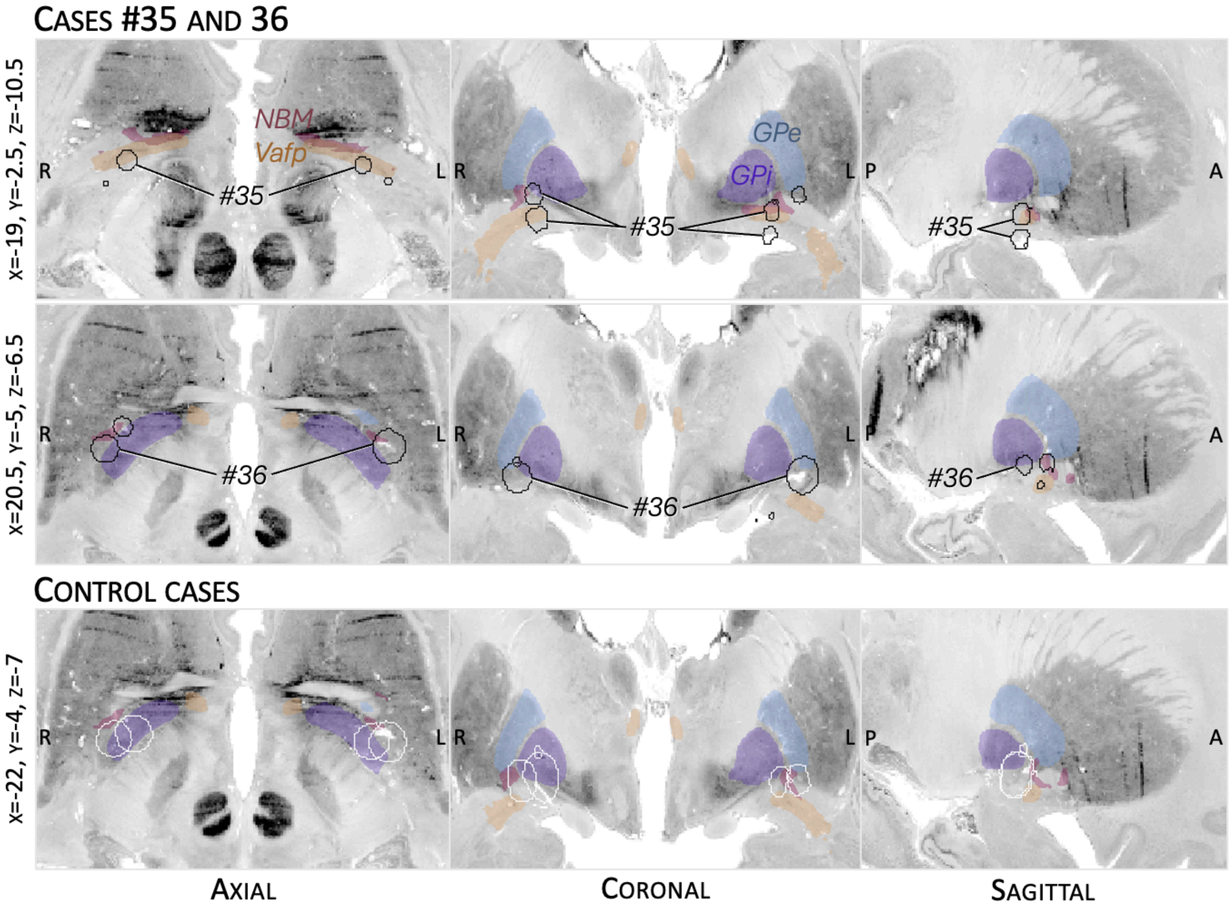


**Figure S11:** Overlap between stimulation volumes (black outlines) and surrounding anatomical structures for Cases #35 and #36 (NBM-DBS, see Table S3 for details). Lower panels show overlap between stimulation volumes (white outlines) and anatomical structures for two control cases from the same study. These two patients also had visual hallucinations at baseline, which did not improve under NBM-DBS (Gratwicke et al., 2018). The relief of hallucinations in Cases #35 and #36 may be hypothesized to relate to stimulation of the NBM itself, although the similar degree of overlap between stimulation volumes and NBM between these cases and control cases argues against this explanation. Indeed, the NBM plays a role in sensory gating (Dürschmid et al., 2020), and the cholinergic system is involved in visual hallucinations in PD (Ignatavicius et al., 2025). NBM-DBS may increase acetylcholine release (Kurosawa et al., 1989; Rasmusson et al., 1992), although this is unlikely to be the main mechanism of DBS, especially in the context of early degeneration of the cholinergic system in PD (Braak et al., 2004). Structures are shown as defined in the DISTAL atlas (Ewert et al., 2018), CIT168 atlas (Pauli et al., 2018), and atlas of the human hypothalamus (Neudorfer et al., 2020). GPe: Globus pallidus externus, GPi: Globus pallidus internus, NBM: Nucleus Basalis of Meynert, Vafp: Ventral amygdalofugal pathway.

| Right VTA | | Left VTA | |
|---|---|---|---|
| *Case #35* | | | |
| CM amygdala | 15.9% - 8.1 $mm^3$ | CM amygdala | 42% - 17.2 $mm^3$ |
| GPi | 10.5% - 5.3 $mm^3$ | NBM | 7.9% - 3.2 $mm^3$ |
| NBM | 10.1% - 5.2 $mm^3$ | | |
| GPe | 1.9% - 1.0 $mm^3$ | | |
| *White matter pathways (bilateral)* | | | |
| Vafp, 90.9% of fibers | | | |
| AC, 66.6% of fibers | | | |
| *Case #36* | | | |
| GPi | 20.2% - 13.9 $mm^3$ | GPe | 17.3% - 18.3$mm^3$ |
| CM amygdala | 14.2% - 9.8 $mm^3$ | NBM | 6.6% - 7.0 $mm^3$ |
| NBM | 9.5% - 6.5 $mm^3$ | GPi | 4.5% - 4.8 $mm^3$ |
| GPe | 4.9% - 3.4 $mm^3$ | | |
| *White matter pathways (bilateral)* | | | |
| AC, 23.1% of fibers | | | |
| *Control case 1* | | | |
| CM amygdala | 35.3% - 64.7 $mm^3$ | GPi | 11.5% - 22.5 $mm^3$ |
| GPi | 32.5% - 59.4 $mm^3$ | GPe | 11.2% - 21.9 $mm^3$ |
| NBM | 2.5% - 4.6 $mm^3$ | CM amygdala | 4.0% - 7.9 $mm^3$ |
| GPe | 1.1% - 2.1 $mm^3$ | NBM | 2.2% - 4.4 $mm^3$ |
| *White matter pathways (bilateral)* | | | |
| Vafp, 5.9% of fibers | | | |
| AC, 4.8% of fibers | | | |
| St, 3.9% of fibers | | | |
| *Control case 2* | | | |
| GPi | 54.7% - 95.3 $mm^3$ | GPi | 27.8% - 49.9 $mm^3$ |
| CM amygdala | 12.5% - 21.8 $mm^3$ | CM amygdala | 13.0% - 23.3 $mm^3$ |
| | | NBM | 2.4% - 4.4 $mm^3$ |
| *White matter pathways (bilateral)* | | | |
| Vafp, 5.1% of fibers | | | |
| St, 1.5% of fibers | | | |

**Table S14:** Overlap between stimulation volumes (“VTAs”) and surrounding anatomical structures and white matter pathways for Cases #35 and #36 (NBM-DBS), as well as for two control cases from the same study (i.e., two patients with visual hallucinations at baseline which did not improve under NBM-DBS, Gratwicke et al., 2018). Structure overlaps are given in percentage of total VTA volume and in $mm^3$ (only structures with overlap of >1% of VTA and pathways with overlap of >1% of fibers are included). Anatomical structures were from the DISTAL atlas (Ewert et al., 2018), Jülich atlas (Amunts et al., 2005) and atlas of the human hypothalamus (Neudorfer et al., 2020). AC: Anterior commissure, GPe: Globus pallidus externus, GPi: Globus pallidus internus, NBM: Nucleus Basalis of Meynert, St: Stria terminalis, Vafp: Ventral amygdalofugal pathway.

### Supplementary material 2: Early attempts at deep brain stimulation for psychosis

Several authors have attempted intracranial brain stimulation in patients with psychosis in the second half of the 20th century.

Sem-Jacobsen performed acute brain stimulation in patients with psychosis in the context of implanting depth electrodes in the ventromedial frontal lobe to perform electrophysiological recordings prior to lobotomy. While these stimulations occasionally elicited relaxation, pleasurable sensations, and –according to some reports– transient improvements in psychotic symptoms, stimulation was not systematically pursued as a therapeutic option at the time (Dietrichs, 2022; C. Sem-Jacobsen, 1959; C. W. Sem-Jacobsen et al., 1956). Delgado also conducted stimulation of the same areas, with similar effects. While his stimulation device allowed for prolonged periods of stimulation, he himself acknowledged that the stimulation had no reliable therapeutic effects (Delgado et al., 1952, 1955). Detailed report of behavioral effects of stimulation could not be retrieved from the literature.

To the best of our knowledge, only Robert Heath pursued chronic deep brain stimulation as a treatment for schizophrenia or psychosis (R. Heath, 1954). It should be mentioned that much of this work raised intense controversy (A. Baumeister, 2011; O'Neal et al., 2017). While Heath always emphasized his therapeutic motivation for the interventions, Baumeister argued that his stimulation studies may better defined as research than practice, and that they fell short of ethical standards even at the time (A. A. Baumeister, 2000). In his 1954 monograph (R. Heath, 1954), Heath reports on 20 patients with schizophrenia who received stimulation of the septal region (1 to 28 stimulations lasting 4s to 40 min each), with electrodes left in place up to 4 days. While he reported that his focus on the septal region came from animal studies suggesting that the brain network for emotion was primarily subcortical (R. G. Heath, 1996), authors later pointed to inconsistencies in his rationale (A. A. Baumeister, 2000). The electrodes were implanted with an "open technique" allowing for direct visualization of the foramen as an anatomical landmark, meaning that important brain damage occurred (including damage of the capsular white matter), and that infections were common. Indeed, 11 patients presented serious complications from the procedure (including seizures, infections, coma, and death) (R. Heath, 1954). Heath acknowledged that the stimulation parameters he used probably caused brain damage, too (peak current up to 40mA, voltage up to 34V, 100Hz, 1ms rectangular pulse, with reversal of polarity added starting from the 9th patients). Because of these and other confounding factors (A. A. Baumeister, 2000; R. Heath, 1954; R. G. Heath & Mickle, 1957; O'Neal et al., 2017), while Heath claimed that 5 patients "markedly improved" and 8 patients "improved", these putative effects can hardly be attributed to the stimulation itself. Interestingly, Heath noted that improvements were mostly seem in flattened affect and disturbed motor behavior (similar to lobotomies), describing patients as more alert, outgoing, friendly, or less depressed, while hallucinations and delusions were the last symptoms to improve (R. Heath, 1954). The open procedure also meant that electrode placement was very unprecise. Furthermore, several electrodes were implanted in each patient, and the individual patient summaries in the 1954 report show that stimulations were also performed at electrodes that were believed to be outside of the septal region (Fig. S12). One should also mention that what Heath referred to as the septal region is large and anatomically heterogeneous, including the septal nuclei, subcallosal gyrus and the most medial aspect of the nucleus accumbens, as well as numerous white matter pathways and structures such as the rostral part of the corpus callosum, diagonal band of Broca, pre-commissural fornix (see (R. Heath, 1954; R. G. Heath, 1996). Overall, the lack of consistency in the conduct and report of the studies complicates the interpretation of this data. A second series of 22 patients was implanted using stereotactic techniques, and the electrodes were left in place for up to two years (R. G. Heath & Mickle, 1957). No therapeutic benefit was observed in this second series, leading Heath himself to suggest that the benefit observed in the first series may have been mostly due to brain damage. Eventually, Heath concluded that septal stimulation did not lead to sustained benefit and abandoned the procedure after 1955 (A. A. Baumeister, 2000; R. G. Heath & Mickle, 1957).

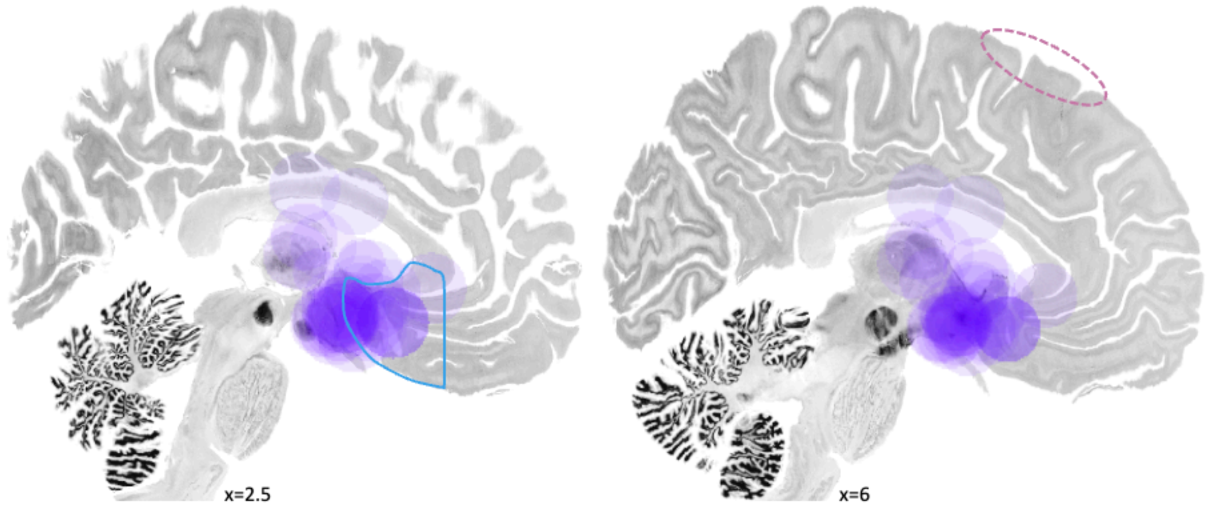


**Figure S12:** Approximative location of stimulation sites in Heath's first series of patients. Locations were derived from descriptions and x-rays available in (R. Heath, 1954). Given the uncertainty associated with the descriptions and information available at the time (x-rays and direct visualization of the foramen of Monro as a landmark during surgery), stimulation sites are shown as 10 mm-radius spheres (purple). Electrodes were implanted in the septal region inferior and anterior to the foramen. In some cases, additional electrodes were implanted in the caudate and thalamus. The electrodes sometimes ended up further away from the intended target. Hence, according to Heath, stimulation sites were putatively in the septal region (blue outline corresponding to his description of this region), hypothalamic or septo-hypothalamic region, caudate nucleus, internal capsule, thalamus and corpus callosum. Pink outline shows approximate location of the skull opening.

| **Hypothesis** | **Proposed follow-up study** |
| --- | --- |
| The two candidate circuits are causally involved in psychotic symptoms | Convergent network mapping: Are lesions causing psychosis and TMS sites improving psychosis functionally or structurally connected to these circuits? |
| Stimulation of the candidate circuits is effective for the treatment of psychosis | Retrospective, quantitative analysis of larger multicenter DBS cohorts (as clinical trials are ongoing internationally): Are stimulation sites more connected to these circuits associated with better clinical outcomes? |
| The mediodorsal nucleus of the thalamus supports reality filtering | Acute stimulation of the mediodorsal thalamus in patients with epilepsy receiving ANT-DBS or implanted with thalamic SEEG electrodes: Does acute electrical stimulation of the MD impacts performance in a reality filtering task? |

**Table S15**: Main hypotheses derived from the present study, and proposed follow-up studies.
